# Global Prevalence of Post-Acute Sequelae of COVID-19 (PASC) or Long COVID: A Meta-Analysis and Systematic Review

**DOI:** 10.1101/2021.11.15.21266377

**Authors:** Chen Chen, Spencer R. Haupert, Lauren Zimmermann, Xu Shi, Lars G. Fritsche, Bhramar Mukherjee

**Affiliations:** Department of Biostatistics, University of Michigan School of Public Health, Ann Arbor, MI 48109, USA; Center for Precision Health Data Science, University of Michigan, Ann Arbor, MI 48109, USA; Rogel Cancer Center, University of Michigan Medicine, Ann Arbor, MI 48109, USA; Center for Statistical Genetics, University of Michigan School of Public Health, Ann Arbor, MI 48109, USA; Department of Epidemiology, University of Michigan School of Public Health, Ann Arbor, MI 48109, USA

## Abstract

**Importance:** As SARS-CoV-2 pervades worldwide, considerable focus has been placed on the longer lasting health effects of the virus on the human host and on the anticipated healthcare needs.

**Objective:** The primary aim of this study is to examine the prevalence of post-acute sequelae of COVID-19 (PASC), commonly known as long COVID, across the world and to assess geographic heterogeneities through a systematic review and meta-analysis. A second aim is to provide prevalence estimates for individual symptoms that have been commonly reported as PASC, based on the existing literature.

**Data Sources:** PubMed, Embase, and iSearch for preprints from medRxiv, bioRxiv, SSRN, and others, were searched on July 5, 2021 with verification extending to August 12, 2021.

**Study Selection:** Studies written in English that consider PASC (indexed as ailments persisting at least 28 days after diagnosis or recovery for SARS-CoV-2 infection) and that examine corresponding prevalence, risk factors, duration, or associated symptoms were included. A total of 40 studies were included with 9 from North America, 1 from South America, 17 from Europe, 11 from Asia, and 2 from other regions.

**Data Extraction and Synthesis:** Data extraction was performed and separately cross-validated on the following data elements: title, journal, authors, date of publication, outcomes, and characteristics related to the study sample and study design. Using a random effects framework for meta-analysis with DerSimonian-Laird pooled inverse-variance weighted estimator, we provide an interval estimate of PASC prevalence, globally, and across regions. This meta-analysis considers variation in PASC prevalence by hospitalization status during the acute phase of infection, duration of symptoms, and specific symptom categories.

**Main Outcomes and Measures:** Prevalence of PASC worldwide and stratified by regions.

**Results:** Global estimated pooled PASC prevalence derived from the estimates presented in 29 studies was 0.43 (95% confidence interval [CI]: 0.35, 0.63), with a higher pooled PASC prevalence estimate of 0.57 (95% CI: 0.45, 0.68), among those hospitalized during the acute phase of infection. Females were estimated to have higher pooled PASC prevalence than males (0.49 [95% CI: 0.35, 0.63] versus 0.37 [95% CI: 0.24, 0.51], respectively). Regional pooled PASC prevalence estimates in descending order were 0.49 (95% CI: 0.21, 0.42) for Asia, 0.44 (95% CI: 0.30, 0.59) for Europe, and 0.30 (95% CI: 0.32, 0.66) for North America. Global pooled PASC prevalence for 30, 60, 90, and 120 days after index test positive date were estimated to be 0.36 (95% CI: 0.25, 0.48), 0.24 (95% CI: 0.13, 0.39), 0.29 (95% CI: 0.12, 0.57) and 0.51 (95% CI: 0.42, 0.59), respectively. Among commonly reported PASC symptoms, fatigue and dyspnea were reported most frequently, with a prevalence of 0.23 (95% CI: 0.13, 0.38) and 0.13 (95% CI: 0.09, 0.19), respectively.

**Conclusions and Relevance:** The findings of this meta-analysis suggest that, worldwide, PASC comprises a significant fraction (0.43 [95% CI: 0.35, 0.63]) of COVID-19 tested positive cases and more than half of hospitalized COVID-19 cases, based on available literature as of August 12, 2021. Geographic differences appear to exist, as lowest to highest PASC prevalence is observed for North America (0.30 [95% CI: 0.32, 0.66]) to Asia (0.49 [95% CI: 0.21, 0.42]). The case-mix across studies, in terms of COVID-19 severity during the acute phase of infection and variation in the clinical definition of PASC, may explain some of these differences. Nonetheless, the health effects of COVID-19 appear to be prolonged and can exert marked stress on the healthcare system, with 237M reported COVID-19 cases worldwide as of October 12, 2021.

**Key Points:** 

**Question:** Among those infected with COVID-19, what is the global and regional prevalence of post-acute sequelae COVID-19 (PASC)?

**Findings:** Globally, the pooled PASC prevalence estimate was 0.43, whereas the pooled PASC prevalence estimate for patients who had to be hospitalized due to COVID-19 was 0.57. Regionally, estimated pooled PASC prevalence from largest to smallest effect size were 0.49 for Asia, 0.44 for Europe, and 0.30 for North America. Global pooled PASC prevalence for 30, 60, 90, and 120 days after index date were estimated to be 0.36, 0.24, 0.29, and 0.51, respectively. Among commonly reported PASC symptoms, fatigue and dyspnea were reported most frequently, with a prevalence of 0.23 and 0.13.

**Meaning:** In follow-up studies of patients with COVID-19 infections, PASC was common both globally and across geographic regions, with studies from Asia reporting the highest prevalence.

## Introduction

Coronavirus Disease 2019 (COVID-19), a highly transmissible disease caused by the severe acute respiratory syndrome coronavirus 2 (SARS-CoV-2), has presented extraordinary challenges to the global healthcare system. Fever, dry cough, fatigue, anosmia, and dyspnea are some of the most common symptoms associated with the acute phase of infection.^1^ Additionally, albeit less commonly, symptoms affecting a wide range of organ systems including the brain, kidney, and heart may also accompany a COVID-19 infection.^2^ With respect to the burden of the virus worldwide, there have been over 237 million COVID-19 cases and over 4.8 million deaths, as of October 12, 2021.^3^ Although the vast majority of those infected survive with an ensuing estimated case fatality rate of 2%, survivors of COVID-19 are known to be at-risk for a variety of sequelae— a condition known as Post-Acute Sequelae of COVID-19 (PASC).^4^ Further obscuring this picture, there is also a large fraction of covert infections due to a multitude of reasons including asymptomatic infections,^5^ barrier to testing^6,7^ and underreporting.^8,9^ Indeed, a recent, extensive review estimated the worldwide pooled asymptomatic percentage of COVID-19 infections to be 35.1% (95% CI: 30.7 to 39.9%), as of August 2021.^10^ Tying to the former, covert infections need be considered with respect to the scope of the prolonged health effects of COVID-19.

In the literature, the occurrence of long-term ailments of COVID-19 appears in a variety of names including PASC, Long COVID, Post-Acute COVID-19 Syndrome (PACS), Chronic COVID-19, and Long Haul COVID-19. It is commonly defined as new or persistent symptoms at 4 or more weeks from infection with SARS-CoV-2.^4^ Carfi et al. were among the first to report post-COVID-19 complications, finding 87.4% of hospitalized patients had at least one persistent symptom at a mean of 60.3 days after symptom onset.^11^ Early in 2021, a large UK-based study found that rates of respiratory disease, diabetes, and cardiovascular disease were 6.0 (95% CI: 5.7, 6.2), 1.5 (95% CI: 1.4, 1.6), and 3.0 (95% CI: 2.7, 3.2) times higher, respectively, in those with a COVID-19 diagnosis as compared to matched controls at a mean follow-up of 140 days.^12^ A more recent meta-analysis estimated 80% of those infected with SARS-CoV-2 develop at least one long-term symptom, with the most prevalent symptoms being fatigue, headache, attention disorder, hair loss, and dyspnea.^13^ However, as the meta-analysis was conducted in the earlier stage of the pandemic, the review was limited by the inherently smaller sample size of infected individuals underlying the existing studies at that time. Upon this base, further research was forged into the potential factors that see increased PASC prevalence.

Time since infection, acute phase severity, geographic region, and select sociodemographic characteristics, such as age and sex, are among the constellation of factors likely to influence PASC prevalence estimates. Although a large proportion of the current evidence focuses on the hospitalized COVID-19 population, a German study found 34.8% of COVID-19 patients, with only a mild acute infection, had PASC at 7 months.^14^ To illustrate the geographic heterogeneity seen in PASC prevalence estimates, specific studies from the USA, Italy, and China report prevalence of 28%, 51%, and 76%, respectively.^15–17^ Regarding demographic factors, Sudre et al. found female sex to be associated with developing PASC.^18^ Although no existing global reviews (at the time of this report) present age-specific PASC prevalence, Nasserie et al. (2021) offer some evidence that prolonged symptoms are not distinct to older versus younger age groups.^19^ That is, although the bulk of PASC exhibiting individuals across the included studies were older (median age near 60 years), younger age groups were also found to comprise a non-negligible number of those with persistent symptoms.^19^ Existing research suggests older age to be associated with a moderate increased risk of persisting symptoms (for ten-year increments past age of 40, estimated odds ratio (OR) is 1.10 [95% CI: 1.01-1.19]).^20^ Moreover, the existing inequities by race/ethnicity as it pertains to PASC remain largely unexplored,^21^ despite the same having been shown for COVID-19. Select comorbidities have been identified as being associated with PASC in the existent literature (e.g., increased risk of PASC among individuals with asthma,^18^ though these findings are generally in early stages).

Following these collective efforts, we too emphasize that PASC must be well-defined and well-understood to enable data to inform clinical decision-making and guidance, and thereby, aid the millions of affected individuals worldwide. At this juncture of being nearly two years into the COVID-19 pandemic, numerous large, high-quality studies on PASC, with substantial follow-up time, have been conducted. Expanding on previous meta-analyses hampered by smaller sample sizes and shorter follow-up times, this systematic review and meta-analysis aims to provide a comprehensive synthesis of information on prevalence and symptoms of PASC to-date. Based on previous research, we hypothesize that PASC is common across geographic and demographic groups, with respiratory, neurological, cardiac, and psychological symptoms having the highest prevalence. We close with some avenues for future research considerations, as highlighted by the findings of this review.

## Methods

### Search Strategy

We employed PICO and PRISMA frameworks to guide our entire research process (**eTable 1**).^22^ The literature databases, PubMed and Embase for published articles, as well as iSearch for preprint articles from bioRxiv, medRxiv, SSRN, Research Square, and preprints.org, were searched on July 5, 2021, and search verification was extended through August 12, 2021. The search aimed to capture papers relating to PASC and that examine prevalence, risk factors, and/or duration, published during the years 2020-2021, and written in English. We adapted some search components from a public resource made available by Yale University Libraries.^23^ The full search strategy, including filters for each database, is presented in **eMethods 1**.

### Screening Procedure

A two-step approach to screening was used with an initial title/abstract screening, followed by a full-text screening, and an ultimate discussion and re-examination to resolve conflicting marks. Screeners 1 and 2 performed both phases of the screening independently (i.e., were blinded). Rayyan, a web-based application, was used as a tool to help expedite literature screening for systematic reviews.^24^

Our inclusion criteria were as follows: (1) human study population with confirmed COVID-19 diagnosis through PCR test, antibody test, or diagnosis, (2) index date of first test/diagnosis, date of hospitalization, discharge date, or date of clinical recovery/negative test, (3) primary outcome must include prevalence, risk factors, duration, or symptoms of PASC, and (4) the follow-up time is at least 28 days after the index date. We excluded case studies, reviews, studies with imaging or molecular/cellular testing as primary results, and studies with only healthcare workers or residents of nursing homes/long-term care facilities. We also excluded studies that did not meet the sample size threshold of 323, pre-calculated herein. The reason for this is to ensure the included studies were adequately powered to achieve a margin of error of at least 0.05 on the provided PASC prevalence estimate. The sample size threshold was calculated with an estimated prevalence of 30% and for a 95% Wald-type confidence interval for binomial proportion; see **eMethods 2** for further details.

### Data Extraction

After studies were selected, the following relevant data elements were manually extracted separately by both screeners 1 and 2: article title, authors, date of publication, study purpose, study design, population, setting, country, sample size, method of COVID-19 confirmation, index date, follow-up time, demographic variables (i.e., age and sex), and outcomes examined. In the instance of multiple study versions with the same underlying population, we used the most recently published article.

### Outcomes and measures

The primary outcome was the prevalence of PASC and symptoms at least 28 days after the index date. We defined PASC as having any symptoms, or at least one new or persisting symptom during the follow-up time. The follow-up time of COVID-19 patients across studies was divided into the following four groups: PASC persisting at 28-30 days (labeled as 30 days), 60 days, 90 days, and 120 days after the index date. We combined similar symptoms into a broader concept. For example, we joined together dyspnea, shortness of breath, and problem of breathing reported in different studies into a broader symptom concept of dyspnea (see **eTable 2**). Studies were classified into the following three groups based on the study population of PASC: (1) studies with non-hospitalized COVID-19 positive individuals, (2) studies with hospitalized COVID-19 positive individuals, (3) studies with all COVID-19 positive individuals (i.e., a case-mix with hospitalized and non-hospitalized individuals). In addition to prevalence, we were also interested in the risk factors for PASC as secondary outcomes.

### Statistical Analysis

Meta-analysis with random effects and generic inverse variance weighting was performed to estimate the prevalence of PASC and symptoms, for outcomes reported in at least five studies. Of further note is that upon examining the distribution of PASC prevalence, we apply a logit transformation to the proportion. The confidence interval was calculated incorporating between-study variance obtained by the DerSimonian-Laird (DL) estimator (**eMethods 3**). Heterogeneity among studies was reflected by the *I*^2^ statistic, where *I*^2^ between 75% and 100% indicates considerable heterogeneity. We further stratified our analysis by (1) study population type (hospitalized versus mixed hospitalized and non-hospitalized), (2) sex (female versus male), (3) follow-up time, (4) region (Asia, Europe, and USA). Another stratified analysis is presented in the supplement (see **eFigure 1**) wherein pooled PASC prevalence is estimated (A) among studies defining PASC to be persisting symptoms (i.e., extended beyond a pre-specified number of days) and (B) among studies defining PASC to be at least one symptom or not recovered from COVID-19. All analyses were conducted in R (version 4.0.2) using packages meta^25,26^ and metafor.^27^

For critical appraisal, we used a checklist-based tool from Joanna Briggs Institute (JBI), corresponding to prevalence studies and hence, enabling assessment of risk of bias among the included study designs.^28^ Assessment of publication bias was carried out visually by generating funnel plot and formally by conducting Egger’s and Begg’s tests for funnel plot asymmetry (for further details, see **eMethods 4** and **eFigure 2**).

## Results

### Search Results

In our main literature search, we identified 4,438 unique citations of which 270 had titles or abstracts that passed our criteria for a full-text assessment. After the full-text screen, we deemed 40 studies eligible for a qualitative synthesis, of which we further meta-analyzed reported measures from 34 with compatible outcomes. See the PRISMA flow diagram (**Figure 1**) and **eTable 3** for details concerning study inclusion/exclusion criteria. In efforts to further verify the search results, we performed a second literature search one month after the first screen, although no additional eligible studies were identified (**eMethods 1** and **eFigure 3**).

**Figure 1.**
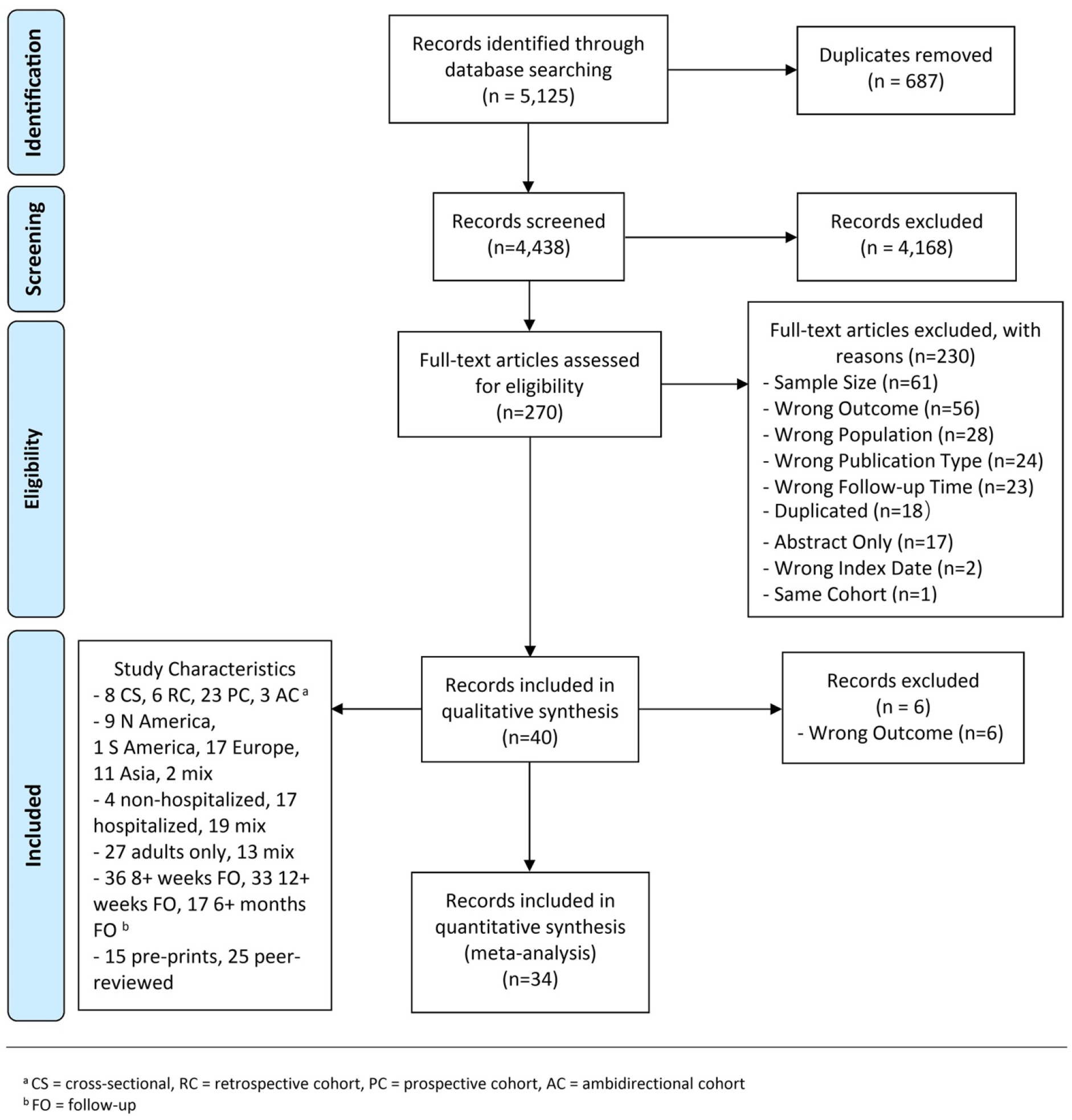
PRISMA flow diagram. *Note*: Additional study characteristics of all included studies are listed in the box in the bottom left.

### Study Characteristics

**Table 1** shows the characteristics of all 40 included articles. The studies comprised a total of 886,388 COVID-19 positive patients that we categorized into non-hospitalized (3,371 patients from 4 studies), hospitalized (61,247 patients from 17 studies), and any COVID-19 positive patients regardless of severity or hospitalization status (821,770 individuals from 19 studies). While only studies with at least 4 weeks follow-up were selected, several studies had data on substantially longer follow-up times: for at least 8 weeks (36 studies), 12 weeks (33 studies), and 6 months (17 studies). **Figure 1** lists additional study characteristics.

**Table 1.** Summary of Included Studies.

| Region | Date of Publication | Authors | Study Design<sup>a</sup> | Population of Interest<sup>b,e</sup> | Setting | Country | Sample Size | Follow-up Time<sup>f</sup> | Age | Sex (% female) | Outcomes of Interest<sup>e</sup> |
| --- | --- | --- | --- | --- | --- | --- | --- | --- | --- | --- | --- |
| Asia | Dec 2020 | Huang et al<sup>29</sup> | AC | COVID-19+, hospitalized adults | Leishenshan Hospital (Wuhan) | China | 464 | 4-6 months *** | 57 (15-93)<sup>c</sup> | 48.50% | Symptom prevalence, risk factors |
|  | Jan 2021 | Huang et al<sup>17</sup> | AC | COVID-19+ adults | Jin Yin-tan Hospital (Wuhan) | China | 1733 | 186 (175-199) days<sup>c</sup> **** | 57 (47-65)<sup>c</sup> | 48% | PASC and symptom prevalence |
|  | Jan 2021 | Xiong et al<sup>30</sup> | PC | COVID-19+, hospitalized adults | Renmin Hospital of Wuhan University | China | 538 | 97 (95-102) days<sup>c</sup> *** | 52 (41-62)<sup>c</sup> | 54.50% | PASC and symptom prevalence |
|  | Jan 2021 | Zheng et al<sup>31</sup> | CS | COVID-19+, hospitalized adults | Multicenter (hospitals in Wuhan) | China | 574 | 241.79 (16.16) days<sup>d</sup> **** | 57.7 (11.4)<sup>d</sup> | 60.60% | Symptom prevalence |
|  | Apr 2021 | Shang et al<sup>32</sup> | PC | COVID-19+, severe, hospitalized | Multicenter (3 hospitals in Wuhan) | China | 796 | 6 months *** | 62 (51-69)<sup>c</sup> | 49.20% | PASC and symptom prevalence, risk factors |
|  | June 2021 | Areekal et al<sup>33</sup> | CS | COVID-19+, hospitalized, symptomatic adults | Government Medical College, Thrissur (Kerala) | India | 335 | 28 days * | 50.7 (15.7)<sup>d</sup> | 48.10% | PASC and symptom prevalence, risk factors |
|  | June 2021 | Budhiraja et al<sup>34</sup> | PC | COVID-19+, hospitalized | Multicenter (3 Hospitals in North India) | India | 990 | 9 (4-12) months<sup>d</sup> **** | 14.6% <=29<br>59.7% 30-59<br>25.7% 60+ | 67.70% | PASC and symptom prevalence |
|  | July 2021 | Naik et al<sup>35</sup> | PC | COVID-19+ adults | Tertiary Care Facility in New Delhi | India | 1,234 | 91 (45-185) days<sup>c</sup> *** | 41.4 (14.2)<sup>d</sup> | 30.60% | PASC and symptom prevalence, risk factors, duration |
|  | Mar 2021 | Mannan et al<sup>36</sup> | CS | COVID-19+, hospitalized | Multicenter (6 hospitals) | Bangladesh | 1,021 | 4+ Weeks * | 1.8% 0-9<br>4.9% 10-19<br>24.4% 20-29<br>30.4% 30-39<br>16.8% 40-49<br>12.4% 50-59<br>9.4% 60+ | 25% | Symptom prevalence |
|  | Nov 2020 | Sami et al<sup>37</sup> | PC | COVID-19+, hospitalized adults | Khorshid Hospital (Isfahan) | Iran | 452 | 4 weeks * | n/a | n/a | Symptom prevalence |
|  | Aug 2021 | Munblit et al<sup>38</sup> | PC | COVID-19+, hospitalized adults | Multicenter (Sechenov University Hospital Network, Moscow) | Russia | 2,649 | 218 (200-236) days<sup>c</sup> **** | 56 (46-66)<sup>c</sup> | 51.10% | PASC and symptom prevalence, risk factors |
| Europe | Jan 2021 | Venturelli et al<sup>16</sup> | PC | COVID-19+ adults | Papa Giovanni XXIII Hospital (Bergamo) | Italy | 767 | 105 (84-127) days<sup>c</sup> *** | 63 (13.6)<sup>d</sup> | 32.90% | PASC and symptom prevalence |
|  | Feb 2021 | Soraas et al<sup>39</sup> | PC | COVID-19+, non-hospitalized adults | Online Survey | Norway | 588 | 248 (18) days<sup>d</sup> **** | 48<sup>d</sup> | 57% | PASC and symptom prevalence |
|  | Mar 2021 | Morin et al<sup>40</sup> | PC | COVID-19+, hospitalized adults | Bicêtre Hospital (Paris) | France | 478 | 113 (94-128) days<sup>c</sup> *** | 61 (16)<sup>d</sup> | 42.10% | PASC and symptom prevalence |
|  | Mar 2021 | Lampl et al<sup>41</sup> | RC | COVID-19+ | Regensburg Public Health Department, Regensburg, Bavaria | Germany | 419 | 6+ weeks * | 44 (30-57)<sup>c</sup> | 56.60% | PASC and symptom prevalence |
|  | Mar 2021 | Ayoubkhani et al<sup>12</sup> | RC | COVID-19+, hospitalized | NHS hospitals in England | UK | 47,780 | 140 (50) days<sup>d ***</sup> | 64.5 (19.2)<sup>d</sup> | 45% | Symptom prevalence |
|  | Apr 2021 | Lemhofer et al<sup>42</sup> | CS | COVID-19+ adults with mild to moderate infection | Survey Administered by 2 Bavarian health departments | Germany | 365 | 3 months+<br>*** | 49.8 (16.9)<sup>d</sup> | 58.50% | PASC and symptom prevalence |
|  | May 2021 | Orrù et al<sup>43</sup> | CS | COVID-19+ adults plus COVID-19- controls | Online Survey (recruitment via social media or email) | Italy | 507 | 1-3 months<br>** | 0.2% <20<br>12.23% 20-29<br>20.91% 30-39<br>30.77% 40-49<br>26.04% 50-59<br>8.24% 60-69<br>1.58% >70 | 82.05% | Symptom prevalence |
|  | May 2021 | Desgranges et al<sup>44</sup> | PC | COVID-19+, symptomatic, outpatient adults with at least one risk factor for severe COVID-19 | University hospital of Lausanne | Switzerland | 418 | 105 (121-204) days<sup>c ***</sup> | 41 (31-54)<sup>c</sup> | 62% | PASC and symptom prevalence, risk factors |
|  | June 2021 | Peghin et al<sup>45</sup> | AC | COVID-19+ adults | Udine Hospital | Italy | 599 | 191 (172-204) days<sup>c ****</sup> | 53(15.8)<sup>d</sup> | 53.40% | PASC and symptom prevalence |
|  | June 2021 | Righi et al<sup>46</sup> | PC | COVID-19+ adults | Verona University Hospital | Italy | 448 | 6 weeks, 12 weeks<br>** | 56 (45-66)<sup>c</sup> | 45.10% | PASC and symptom prevalence |
|  | June 2021 | Maestre-Muñiz et al<sup>47</sup> | CS | COVID-19+ adults | Tomelloso General Hospital | Spain | 543 | 12 months<br>**** | n/a | n/a | PASC and symptom prevalence |
|  | July 2021 | Ghosn et al<sup>48</sup> | PC | COVID-19+, hospitalized | Multicenter (French Covid Cohort) | France | 1,137 | 3 months, 6 months<br>*** | 61 (51-71)<sup>c</sup> | 37% | Symptom prevalence |
|  | July 2021 | Augustin et al<sup>14</sup> | PC | COVID-19+, non-hospitalized adults | University Hospital Cologne | Germany | 1.4 months: 958<br>4.3 months: 442<br>6.8 months: 353 | 1.4 (1-2) months,<br>4.3 (3-5) months,<br>6.8 (6-8) months<sup>c</sup><br>**** | 43 (31-54)<sup>c</sup> | 53.50% | PASC and symptom prevalence, risk factors |
|  | July 2021 | Menges et al<sup>49</sup> | PC | COVID-19+ adults | Department of Health of the Canton of Zurich, Switzerland Surveillance | Switzerland | 431 | 7.2 (5.9-10.3) months<sup>c</sup><br>**** | 47 (33-58)<sup>c</sup> | 50% | PASC and symptom prevalence, risk factors |
|  | July 2021 | Taylor et al<sup>50</sup> | PC | COVID-19+, hospitalized | Barts Health NHS Trust (London) | UK | 675 | 12+ weeks<br>*** | n/a | 42.10% | Symptom prevalence |
|  | Aug 2021 | Fernández-de-Las-Peñas et al<sup>51</sup> | PC | COVID-19+, hospitalized | Multicenter | Spain | 1,142 | 7 (0.6) months<sup>d</sup><br>**** | 61 (17)<sup>d</sup> | 48% | PASC and symptom prevalence |
|  | Apr 2021 | Whittaker et al<sup>52</sup> | PC | COVID-19+ adults | Clinical Practice Research Database (CPRD) Aurum | UK | 46,687 | 63 days (63-63)<sup>c **</sup> | 38.6% 18-30<br>16.6% 31-40<br>15.7% 41-50<br>16% 51-60<br>7.4% 61-70<br>3.3% 71-80<br>2.4% >80 | 54.60% | Symptom prevalence |
| Americas | Dec 2020 | Cirulli et al<sup>53</sup> | PC | COVID-19+ adults | Surveyed participants from Helix DNA Discovery Project and Healthy Nevada Project | USA | 357 | 30, 60, 90 days *** | n/a | n/a | PASC and symptom prevalence, risk factors |
|  | Mar 2021 | Hirschtick et al<sup>54</sup> | CS | COVID-19+, symptomatic adults | Michigan Disease Surveillance System | USA | 593 | 30, 60 days ** | 51.5 (15.8)<sup>d</sup> | 56.10% | PASC prevalence |
|  | Mar 2021 | Spotnitz et al<sup>15</sup> | RC | COVID-19+ | ICM MarketScan Commercial Claims and Encounters, Optum Electronic Health Record, and Columbia University Irving Medical Center. | USA | 448,176 | 30-180 days *** | n/a | n/a | PASC and symptom prevalence |
|  | Mar 2021 | Perlis et al<sup>20</sup> | CS | COVID-19+, symptomatic | Online Survey with Non-Probability Sampling | USA | 6,211 | 10 months **** | 37.8 (12.2)<sup>d</sup> | 45.10% | PASC and symptom prevalence, risk factors |
|  | Mar 2021 | Huang et al<sup>55</sup> | RC | COVID-19+, non-hospitalized with 5+ year history in EHR system | UC CORDS (University of California Covid research data set) | USA | 1,407 | 61+ days ** | 2% < 18<br>10% 18-29<br>16% 30-39<br>18% 40-49<br>21% 50-59<br>16% 60-69<br>12% 70-79<br>6% >= 80 | 58.90% | Symptom prevalence, risk factors |
|  | Apr 2021 | Damiano et al<sup>56</sup> | PC | COVID-19+, hospitalized (moderate or severe Covid) adults | Hospital das Clínicas da Faculdade de Medicina da Universidade de São Paulo | Brazil | 425 | 207 (20.4) days<sup>d</sup> **** | 55.7 (14.2)<sup>d</sup> | 51.53% | Psychiatric and cognitive symptom prevalence |
|  | Apr 2021 | Chevinsky et al<sup>57</sup> | PC | COVID-19+ adult inpatients and outpatients | Premier Healthcare Database Sepical COVID-19 Release (PHD-SR) | USA | Outpatients: 44,489<br>Inpatients: 27,284 | 31-60 days, 61-90 days, 91-120 days *** | Inpatients: 8.8% 18-39<br>10% 40-49<br>28.1% 50-64<br>22.1% 65-74<br>18% 75-84<br>13% >= 85<br>Outpatients: 35.7% 19-39<br>18.1% 40-49<br>25.9% 50-64<br>10.3% 65-74<br>6% 75-84<br>4% >= 85 | 52.5% (inpatients)<br>61.2% (outpatients) | PASC and symptom prevalence |
|  | May 2021 | Yomogida et al<sup>58</sup> | PC | COVID-19+ adults | Long Beach Department of Health and Human Services Surveillance | USA | 366 | 1 Month, 2 months, 10 weeks-5 months *** | 11.4% 18-24<br>39.3% 25-39<br>30.2% 40-54<br>10.6% 55-64<br>8.2% 65+ | 56.40% | PASC and symptom prevalence, risk factors |
|  | June 2021 | Wong-Chew et al<sup>59</sup> | PC | COVID-19+, hospitalized adults | Temporary Covid-19 Hospiital in Mexico City | Mexico | 30 days: 1,303<br>90 days: 928 | 30, 90 days *** | n/a | n/a | Symptom prevalence, risk factors |
|  | June 2021 | Shoucri et al<sup>60</sup> | RC | COVID-19+, hospitalized adults | New York-Presbyterian/Columbia University Irving Medical Center | USA | 3 months: 488<br>6 months: 364 | 3, 6 months *** | 3 months: 60 (47.8-71)<br>6 months: 61 (50.0-71) <sup>c</sup> | 43.2% - 3 months<br>47.8% - 6 months | Symptom prevalence |
| Mix | Mar 2021 | Sudre et al<sup>18</sup> | PC | COVID-19+, symptomatic | COVID Symptom Study App | UK, Sweden, US | 4,182 | 28-84 days ** | 42 (32-53) <sup>c</sup> | 71.50% | PASC and symptom prevalence, risk factors, duration |
|  | May 2021 | Taquet et al<sup>61</sup> | RC | COVID-19+, age 10+ | TriNetX EHR Network | USA, others | 236,379 | 6 months *** | 46 (19.7) <sup>d</sup> | 55.60% | Risk factors, duration |

<sup>a</sup> PS = Prospective Cohort, RS = Retrospective Cohort, CS = Cross-sectional, AC = Ambidirectional Cohort
<sup>b</sup> Not all inclusion/exclusion criteria listed
<sup>c</sup> Median (IQR) or Median (range)
<sup>d</sup> Mean (SD) or Mean (95% CI)
<sup>e</sup> Some studies included populations and outcomes outside the scope of this review.
<sup>f</sup> Asterisks denote length of follow-up time according to the following convention: [4 weeks, 8 weeks] - \*, (8 weeks, 12 weeks] - \*\*, (12 weeks, 6 months] - \*\*\* 6+ months - \*\*\*\*. If a study considered measurements at several follow-up times, the longest duration was used.

### PASC Prevalence

Among the 34 included studies in the quantitative synthesis, we meta-analyzed the 29 studies reporting an overall prevalence of PASC. Pooled global PASC prevalence was estimated to be 0.43 (95% CI: 0.35, 0.63) (**Table 2**). Substantial heterogeneity was observed among the included studies (*I*^2^ = 100%, P < 0.001). Estimates ranged widely from 0.09 to 0.81 which may in part be driven by differences in terms of sex, region, COVID-19 study population, and follow-up time. For example, the studies that included only hospitalized cases tended to show higher PASC prevalence than non-hospitalized or the mix of hospitalized and non-hospitalized patients (**Figure 2**). To better understand the interplay of these factors with PASC prevalence estimates, we performed additional stratified meta-analyses (**Table 2**).

**Figure 2.**
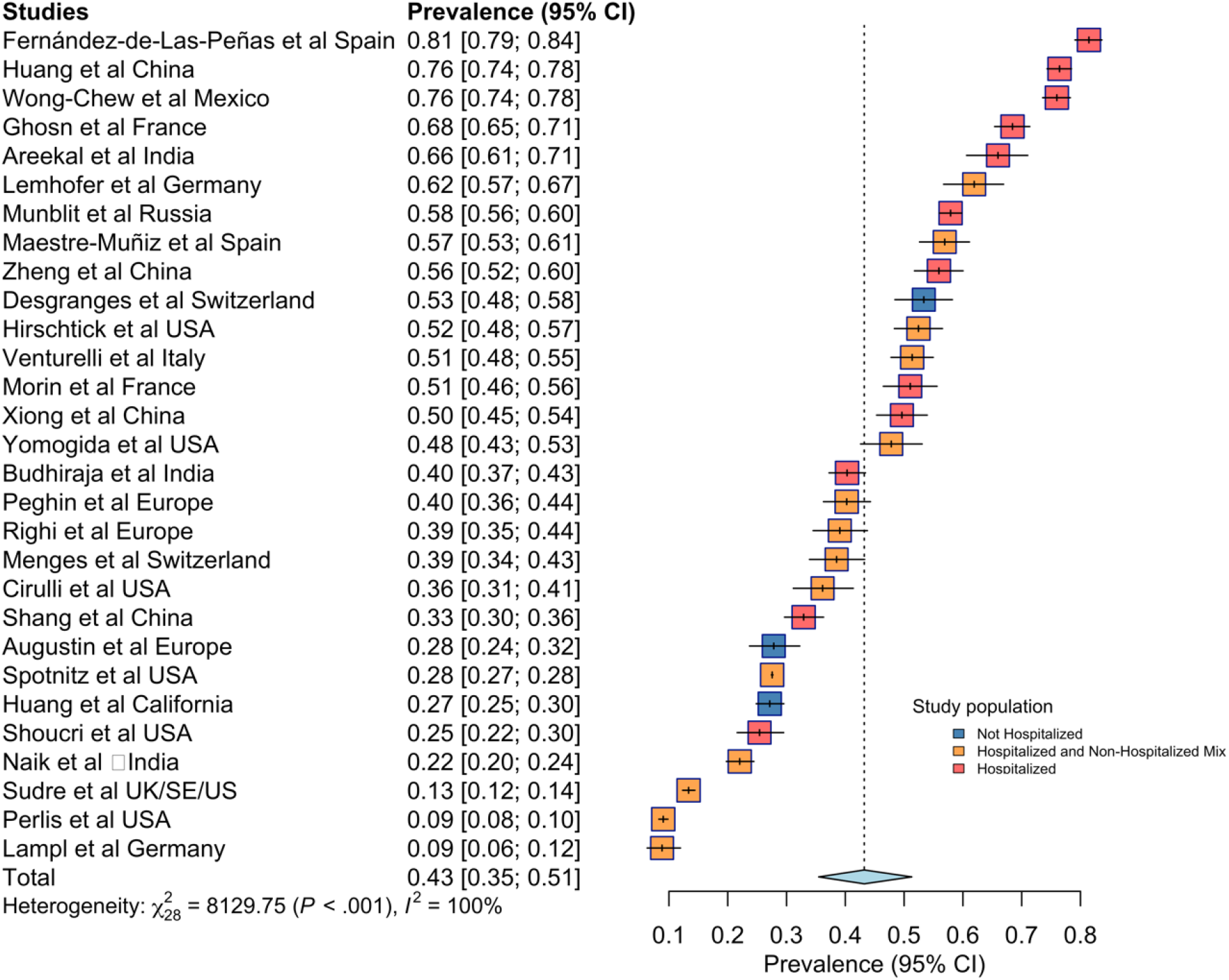
Forest plot for worldwide PASC prevalence. *Notes*: Prevalence estimates and 95% CIs are provided for each study with a relevant measure, and for the meta-analysis of all such studies. For individual studies, the horizontal line represents the estimate, whiskers represent the confidence interval, the size of the box represents the weight assigned to the study, and the color shading reflects the hospitalization status of the study population, as noted in the legend. For the pooled estimate, the width of the diamond represents the confidence interval. Meta-analyzed prevalence and 95% CIs are calculated using random-effects models with inverse variance weighting as described in the methods. Measures of heterogeneity of prevalence estimates are provided.

**Table 2.** Meta-analysis of pooled PASC prevalence with 95% CI in COVID-19 Positive Individuals ^a^

|  |  | **PASC Prevalence in COVID-19 Positive Individuals [95% CI];**<br>**(number of included studies)** |  |  |
| --- | --- | --- | --- | --- |
|  |  | **Any\*** | **Mix of Hospitalized &**<br>**Non-Hospitalized** | **Hospitalized** |
| **Overall** |  | 0.43 [0.35; 0.51]; (29) | 0.33 [0.26; 0.42]; (14) | 0.57 [0.47; 0.66]; (12) |
| **Sex** | Female | 0.49 [0.35; 0.63]; (9) | ... | ... |
|  | Male | 0.37 [0.24; 0.51]; (9) | ... | ... |
| **Region** | Europe | 0.43 [0.29; 0.58]; (13) | ... | ... |
|  | Asia | 0.49 [0.32; 0.66]; (7) | ... | ... |
|  | USA | 0.30 [0.21; 0.42]; (7) | ... | ... |
| **Follow-up time** | 30 days | 0.36 [0.25; 0.48]; (10) | ... | ... |
|  | 60 days | 0.24 [0.13; 0.39]; (9) | ... | ... |
|  | 90 days | 0.32 [0.14; 0.57]; (9) | ... | ... |
|  | 120 days | 0.51 [0.41; 0.61]; (12) | 0.45 [0.34; 0.57]; (3) <sup>b</sup> | 0.56 [0.44; 0.68]; (8) |
| **General symptoms** | Fatigue | 0.23 [0.13; 0.38]; (22) | 0.22 [0.09; 0.44]; (11) | 0.26 [0.17; 0.38]; (8) |
|  | Tachycardia | 0.07 [0.03; 0.18]; (6) | ... | ... |
|  | Appetite/<br>Eating disorder | 0.06 [0.03; 0.1]; (8) | ... | 0.04 [0.02; 0.08]; (6) |
|  | Dizziness | 0.06 [0.03; 0.13]; (5) | ... | ... |
|  | Sore throat | 0.03 [0.02; 0.05]; (10) | ... | 0.03 [0.01; 0.09]; (5) |
|  | Fever | 0.02 [0.01; 0.04]; (12) | 0.02 [0.01; 0.04]; (6) | ... |
| **Neurologic symptoms** | Sleep problems | 0.13 [0.06; 0.28]; (14) | ... | 0.16 [0.11; 0.23]; (9) |
|  | Memory problems | 0.13 [0.1; 0.18]; (10) | ... | 0.12 [0.09; 0.17]; (7) |
|  | Concentration/<br>Confusion / Brain fog | 0.09 [0.05; 0.17]; (11) | ... | 0.06 [0.03; 0.13]; (6) |
|  | Smell | 0.08 [0.05; 0.12]; (13) | 0.14 [0.06; 0.28]; (5) | 0.05 [0.03; 0.08]; (7) |
|  | Taste | 0.08 [0.04; 0.13]; (10) | ... | 0.04 [0.02; 0.07]; (5) |
|  | Headache | 0.05 [0.03; 0.08]; (17) | 0.03 [0.01; 0.1]; (8) | 0.05 [0.02; 0.11]; (7) |
|  | Smell or Taste | 0.05 [0.01; 0.21]; (9) | 0.04 [0.01; 0.23]; (5) | ... |
| **Respiratory symptoms** | Dyspnea | 0.13 [0.09; 0.19]; (24) | 0.12 [0.07; 0.2]; (13) | 0.16 [0.13; 0.21]; (8) |
|  | Cough | 0.07 [0.05; 0.09]; (24) | 0.06 [0.04; 0.09]; (11) | 0.07 [0.05; 0.12]; (10) |
|  | Chest pain | 0.05 [0.04; 0.07]; (15) | 0.03 [0.02; 0.07]; (6) | 0.07 [0.05; 0.1]; (9) |
| **Psychological symptoms** | Anxiety | 0.10 [0.06; 0.16]; (10) | 0.12 [0.05; 0.25]; (6) | ... |
|  | Depression | 0.10 [0.05; 0.21]; (7) | 0.13 [0.04; 0.34]; (5) | ... |
| **Musculoskeletal symptoms** | Joint pain | 0.13 [0.05; 0.29]; (5) | ... | ... |
|  | Myalgia | 0.06 [0.03; 0.13]; (17) | 0.06 [0.02; 0.16]; (10) | 0.07 [0.04; 0.12]; (7) |
| **Gastrointestinal symptoms** | Abdominal pain | 0.04 [0.02; 0.12]; (6) | ... | ... |
|  | Diarrhea | 0.03 [0.01; 0.07]; (9) | ... | 0.02 [0.01; 0.04]; (5) |
| **Dermatologic symptoms** | Hair loss | 0.07 [0.02; 0.24]; (10) | ... | 0.13 [0.08; 0.21]; (6) |

\* Includes studies that reported on only non-hospitalized, mix of hospitalized & non-hospitalized, or only hospitalized COVID-19 positive patients
<sup>a</sup> Pooled estimates and 95% CIs calculated from random-effect models with inverse variance weighting as described in methods. Prevalence is stratified by acute-phase hospitalization status. Estimates for the non-hospitalized population are not provided due to lack of sample size.
<sup>b</sup> Only 3 studies with mixed hospitalized and non-hospitalized population. This estimate should be interpreted with caution due to low sample size.

First, the pooled PASC prevalence in hospitalized patients of 0.57 (95% CI: 0.45, 0.68) compared to the estimate in a mix of hospitalized and non-hospitalized COVID-19 patients of 0.31 (95% CI: 0.24, 0.40) revealed a sizeable difference, further distinguished by non-overlapping confidence intervals. However, we note that a wide range of estimates contributed to both groups (i.e., PASC prevalence varied from 0.25 – 0.81 in hospitalized studies versus 0.09 – 0.62 in the mixed group, and significant heterogeneity was present in both) (**eFigure 4B**). Next, when focusing on sex, we estimated a pooled PASC prevalence in females of 0.49 (95% CI: 0.35, 0.63), which was higher than that in males of 0.37 (95% CI: 0.24, 0.51). Considering the same studies underly both strata, this imbalance was unlikely attributable to differences in the contributing studies (**eFigure 4A**).

Examining region-specific prevalences, pooled estimated prevalence of PASC was lower in the USA at 0.30 (95% CI: 0.21, 0.42) than in Europe at 0.43 (95% CI: 0.29, 0.58), while the highest estimated prevalence was in Asia at 0.49 (95% CI: 0.32, 0.66). Considerable within-region variation was observed among the included studies in that the corresponding ranges of prevalence of PASC were generally wide, with Europe exhibiting the largest range of 0.09 – 0.81. Overall, we did not identify any patterns with respect to particular countries that could explain the heterogeneity within each of the meta-analyzed regions (P < 0.001; **eFigure 4C**).

Finally, we focused on estimating PASC prevalence stratified by follow-up time. With increasing follow-up time from 30 to 60 days after the index date, the estimated pooled prevalence of PASC decreased from 0.36 (95% CI: 0.25, 0.48) to 0.24 (95% CI: 0.13, 0.39). Pooled prevalence of PASC 90 and 120 days after the index date further increased to 0.29 (95% CI: 0.12, 0.57) and to 0.51 (95% CI: 0.42, 0.59), respectively (**eFigure 4D**). A possible reason for this comparatively high PASC prevalence at 120 days of follow-up time is that the bulk of the studies underlying this estimate concentrated on hospitalized populations (**eFigure 5**). Studies are also likely to experience higher drop-out rates as follow-up time increases, resulting in individuals no longer experiencing symptoms being underrepresented at the later time points. Some studies that measured PASC prevalence at multiple time points experienced a similar phenomenon.^14^

Significant levels of heterogeneity being present within each stratified meta-analysis corroborates that no single factor alone may account for the variation in PASC prevalence, but that rather a combination of factors should be considered. Concerning a perceived ordering of influence, the factors that seem to have the largest bearing on the PASC prevalence (as ordered from highest to lowest) were study population, region, and follow-up time. Furthermore, inconsistent PASC definition is a source of heterogeneity. As detailed in **eFigure 1**, studies measuring PASC as having at least one persistent symptom had lower heterogeneity (as measured by a chi-square test statistic) compared to those measuring PASC as at least one symptom (with symptoms not necessarily starting during the acute phase). Noting that the prevalence of each symptom varied, effect size of PASC prevalence estimates may differ in part due to the underlying symptoms assessed therein. An additional meta-analysis of studies with at least 120 days follow-up stratified by COVID-19 population resulted in similar observed heterogeneity (**eFigure 5**). Ultimately, these findings suggest that such variation may be indelible, as key considerations, such as the definition of PASC itself, as well as other clinical and methodological subcomponents, remain largely in flux.^62^

### Prevalence of specific PASC symptoms

Considering a unified definition of PASC remains under investigation (as discussed in the ***Introduction*** section), it was important to understand the prevalence of specific symptoms after COVID-19. In total, we assessed 23 symptoms reported across 30 studies (**Table 2**, **Figure 3**). The five most prevalent symptoms were the following, with corresponding estimated pooled symptom-specific prevalence: fatigue at 0.23 (95% CI: 0.13, 0.38), dyspnea at 0.13 (95% CI: 0.09, 0.19), insomnia at 0.13 (95% CI: 0.06, 0.28), joint pain at 0.13 (95% CI: 0.05, 0.29), and memory problems at 0.13 (95% CI: 0.10, 0.18). Forest plots for symptom-specific prevalence estimates are presented in **eFigure 6**. We note that a study by Orrū et al. from Italy tended to fall toward the higher end of the observed range for several symptom categories, and as such this outlying study (relative to the other underlying studies) may have skewed the resulting point estimates and confidence intervals to a degree.^43^

**Figure 3.**
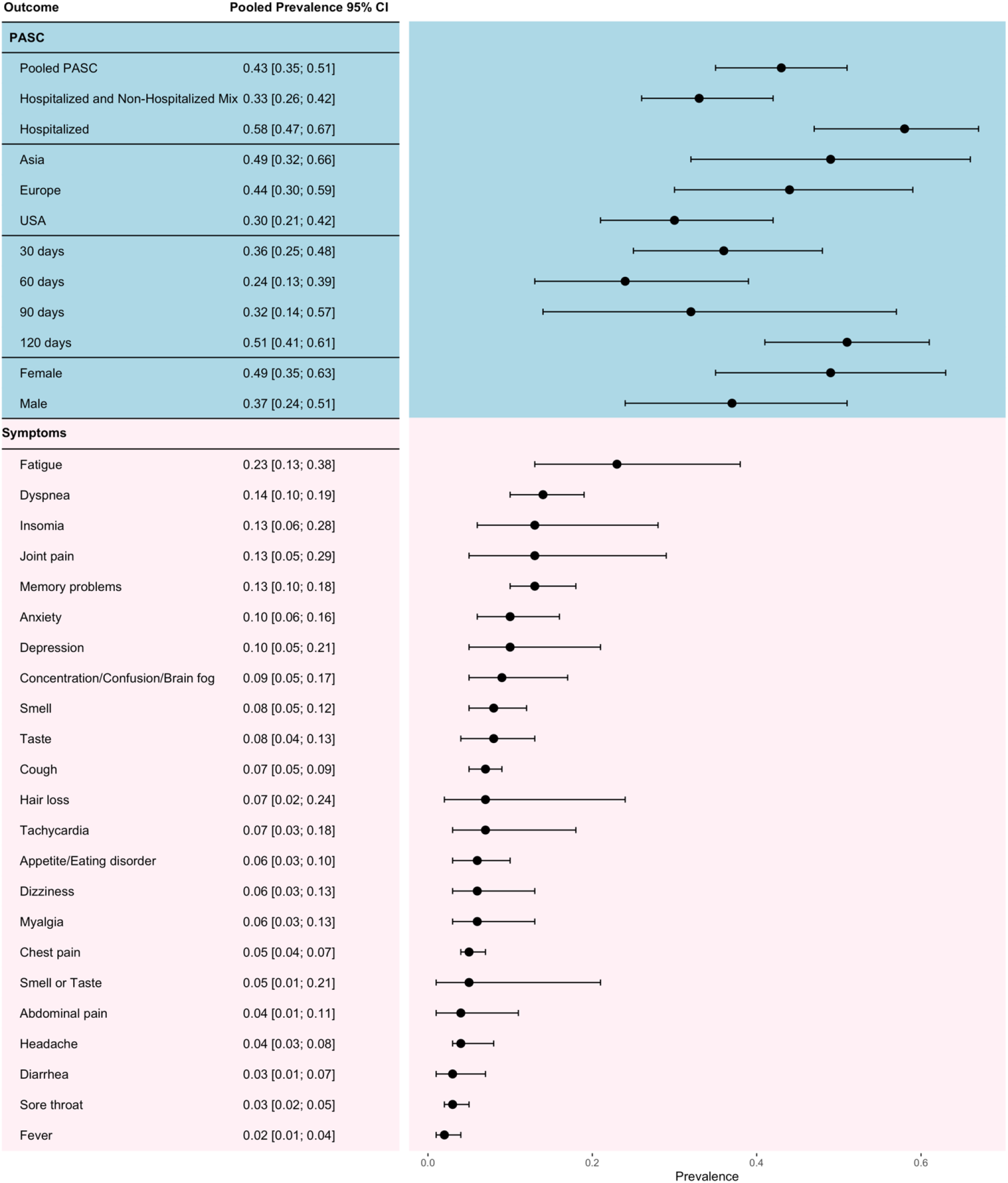
Forest plot for PASC prevalence by hospitalization status, region, follow-up time, and sex, as well as symptom-specific prevalence. *Notes*: Pooled estimates and 95% CIs calculated from random-effect models with inverse variance weighting as described in methods. Pooled estimates with confidence intervals are provided on the left, and visualization of the intervals on the right.

### PASC risk factors

Although all included studies were screened for reported PASC risk factors, sex and pre-existing asthma were the only risk factors that were estimated in multiple studies and thus meta-analyzed. Female sex and pre-existing asthma had higher odds of having PASC with pooled estimated odds ratios (OR) of 1.57 (95% CI: 1.09, 2.26) and 2.15 (95% CI: 1.14, 4.05), respectively. Both meta-analyzed ORs were based on less than 5 studies and should thus be interpreted with caution. Among the studies that were not meta-analyzed, several found that individuals with more severe COVID-19 during the acute phase had higher risk of developing PASC.^37,45,50,52^ Additionally, two studies found older age to be associated with PASC.^20,58^ Other risk factors for PASC including number of symptoms during acute COVID-19,^16^ fatigue^18^, dyspnea,^18,39^ muscle pain,^50^ headache,^18,20^ myalgia,^18^ and pre-existing conditions such as obesity,^18,52^ comorbidity,^45^ and hypothyroidism^37^ were found positively associated with PASC (**eTable 4**).

### Systematic review

Six studies were not included in the meta-analysis since they did not report a composite binary endpoint as prevalence (**Figure 1**). Three studies used incidence rate or incidence density to measure PASC. Chevinsky et al. reported a 7% incidence rate of at least one of the five most common new conditions during days 31 to 120 for inpatients and a 7.7% incidence rate for at least one of 10 new conditions.^57^ A UK study found breathlessness (85 and 536 events per 100,000 person-years in non-hospitalized and hospitalized patients) and joint pain (168 and 295 events per 100,000 person-years in non-hospitalized and hospitalized patients) to be the most common sequelae at 2 months.^52^ Another UK study found the rates of respiratory disease and major cardiovascular events to be 770.5 (95% CI: 757.8, 783.3) and 126 (95% CI: 121, 131) events per 1,000 person-years.^12^ The other three studies investigated PASC with a focus on the psychiatric and neurological illness. Damiano et al. focused on psychiatric and cognitive sequela and reported a prevalence of 0.08 (95% CI: 0.06, 0.11), 0.14 (95% CI: 0.11, 0.18), and 0.16 (95% CI: 0.12, 19) for depression, generalized anxiety disorder, and mixed anxiety-depression.^56^ Another study by Taquet et al. also concentrated on psychiatric disorder and measured incidence and hazard ratio of psychiatric disorder, dementia and insomnia with 90 days follow up.^61^ The estimated probability of having new psychiatric illness 90 days after COVID-19 diagnosis was 5.8% (95% CI: 5.2, 6.4) Huang et al.’s study from China showed psychosocial problems (57.7%), worse depression (35%), and worse dyspnea (32.6%) to be among the most common complaints 4–6 months after discharge.^29^

Two articles described the duration of PASC and persistent symptoms. According to Sudre et al., the median duration of PASC with persisting symptoms was 41 days. For persisting symptoms that occurred at least 28 days after COVID-19 diagnosis, the median duration of persisting fatigue, headache, dyspnea, and myalgia was 33 days, 22 days, 24 days, and 7 days, respectively.^18^ However, the median duration of persisting symptoms was longer in another India study.^35^ A summary table of duration was reported in **eTable 5**.

## Discussion

We screened nearly 4.5 thousand articles and synthesized information from 40 large studies including almost one million individuals worldwide. The empirical findings suggest a global PASC prevalence of approximately 43%. Based on a WHO estimate of 237 million worldwide COVID-19 infections, this global pooled PASC estimate indicates that around 100 million individuals currently experience or have previously experienced long-term health-related consequences of COVID-19. Individuals who were hospitalized during acute COVID-19 infection had higher PASC prevalence at 57%. Female adults had both higher prevalence and risk of having PASC than male adults (49% vs 37%). The prevalence of PASC in Asia, Europe, and USA are approximately 49%, 43%, and 30%, respectively. Next, we contextualize our results among findings from other PASC-related reviews.

Our global PASC estimate of 43% is considerably lower than the 80% figure provided by Lopez-Leon et al.^63^ Their most prevalent sequela was fatigue at 58% which is concordant with fatigue being the most prevalent sequela at 23% in this study. In general, empirical symptom-specific prevalence estimates are lower in this study, although multiple estimates (e.g., for insomnia, memory problems, anxiety, depression) generally reconcile with the Lopez-Leon et al. review.^13^ Similarly, when comparing to the Iqbal et al.^64^ meta-analyzed PASC-related symptom prevalence findings, the estimates herein are lower. A potential reasoning for this is the sample size threshold that we employed may have led to select studies being excluded that were conducted in early 2020 with smaller samples and focused mainly on sicker patients.

Additional notable studies have been published after the date of this systematic search (August 12, 2021), and as such are not captured in the empirical estimates presented herein. As examples, another study by Taquet et al., using the TriNetX Analytics EHR network, estimated 36.55% of COVID-19 patients to have at least one PASC-related symptom 3-6 months after diagnosis.^65^ Huang et al. 2021 also provided an update on their cohort from Jin Yin-tan Hospital in Wuhan.^66^ Their 6-month study (which is included in this review) estimated 6-month PASC to be 76%. Their 12-month update found that, among patients who attended both the 6-month and 12-month follow-up, PASC prevalence decreased from 68% at 6 months to 49% at 12 months.

In addition to experiencing PASC symptoms, some COVID-19 survivors also go on to develop other complications. For the purposes of this review, we define COVID-19 complication as any secondary disease that manifests after the acute phase of a COVID-19 infection. Multisystem Inflammatory Syndrome in Children (MIS-C), Chronic Kidney Disease (CKD), myocarditis/pericarditis, Chronic Fatigue Syndrome (CFS) or myalgic encephalomyelitis, and Kawasaki disease are complications known to be associated with COVID-19.^67^ While the focus of this review is on PASC symptoms rather than complications, further research is necessary to understand the relationship between COVID-19 and these complications and the needs of those living with complications.

Our meta-analysis showed that female sex and pre-existing asthma correspond with higher proportions of PASC development. Outside of meta-analysis, we also found age, acute phase symptoms and severity, hypothyroidism, obesity, hypertension, and other pre-existing conditions to be risk factors for PASC. Protective factors for PASC may also exist, as a recent study suggested vaccines may offer protection.^68^ However, a large hospital-based study suggests the opposite.^69^ As such, the interplay between COVID-19 vaccines and PASC is at-large yet to be determined. Multiple other risk factors for PASC have been detected, and, although encompassed among select included studies, such factors were not meta-analyzed because they did not reach the threshold of at least 5 studies. Increased number of acute-phase symptoms is associated with PASC; however, one study reported 32% of individuals with PASC were asymptomatic during the acute phase in a non-hospitalized population.^55^ Similarly, few studies examined the duration of PASC. Future research needs to further explore risk factors and duration for PASC, as these are generally critical components for clinicians in screening patients for increased risk of developing PASC, and in devising an appropriate treatment protocol accordingly. This leads to the several limitations of this systematic review and meta- analysis.

## Limitations

First, we did not survey grey literature (literature not published in academic journals), which could make our results reflect the positive publication bias known to exist in peer-reviewed medical literature, though we included preprints.^70^ Second, we only considered studies written in English which may have excluded important studies written in other languages. Third, while our criteria for follow-up time and index date seem reasonable, there may be important results from studies using other criteria. For example, a large Danish cohort analyzed by Lund et al. was excluded for their choice of follow-up time.^71^ Fourth, bias in testing for COVID-19, especially in the early stages of the pandemic, might have affected the characteristics of the COVID-19 positive cohort.^72^ In other words, patients without access to testing, patients without strong health-seeking behavior, and asymptomatic individuals are not blanketly reflected in the empirical findings. Additionally, included studies conducted in early 2020 may tend to be older and higher risk individuals, as testing among these groups was prioritized at that time. Fifth, our sample size criteria may have curtailed inclusion of early-pandemic studies, as sample sizes were generally smaller at that time, and thus favored studies examining acute-phase manifestations over studies focusing on PASC. Lastly, while our review included studies across 17+ countries, data from multiple regions are largely absent (notably Africa and Australia). Existing inequities in healthcare access may hamper underserved populations being adequately reflected herein. Moreover, we emphasize that stratifying PASC by race-ethnicity is a noteworthy gap in the literature. With respect to the age composition of the included articles, few children were included in the underlying sample. Future investigators may seek to further examine differences in PASC prevalence among such demographic subgroups.

## Conclusions

Findings from this study provide insight into the empirical estimates of prevalence, symptoms, risk factors, and duration of PASC, with an examination of differences by several factors including geography. We recommend continued attention be focused on identifying patients at-risk of developing PASC and on quantifying duration of PASC to aid in the clinical advancements globally for alleviating the long-lasting health effects of COVID-19.

## Data Availability

All data compiled in the present study are contained in the manuscript.

## Acknowledgements

The authors thank the librarians from the University of Michigan Taubman Health Sciences Library, and in particular, Judith E. Smith, for their guidance on constructing the search strategy for this systematic review.

## Author Contributions

Conceptualization: XS, LGF, BM

Methodology: XS, LGF, BM

Investigation: CC (Screener 1), SRH (Screener 2), XS, LGF, BM

Supervision: XS, LGF, BM

Writing – original draft: CC, SRH, LZ, XS, LGF, BM

Writing – review & editing: CC, SRH, LZ, XS, LGF, BM

## Supplementary Online Content

**eMethods 1.** Systematic review procedure

To perform the systematic review presented herein, we collected both publications and preprints systematically that concern prevalence, risk factors, and/or duration of PASC in any country worldwide. In identifying relevant articles, we searched the following three databases: PubMed, Embase, and iSearch for preprints (encompassing bioRxiv, medRxiv, preprints.org, Research Square, SSRN). The search was conducted on July 5, 2021. Hence, the resulting captured studies reflect those available from January 1, 2020 to July 5, 2021. A second search was conducted on August 12, 2021, to ensure we would not fail to capture recent, important studies. In this search, we used the same search terms and filters as our July 5 search and restricted our attention to studies published in well-established medical journals (i.e., JAMA, Lancet, NEJM, Nature, BMJ, and PloS). Details of study inclusion from the August 12 search can be found in **eFigure 3**. Upon securing citations from the search engines into Mendeley^1^, the reference manager, the resulting body of citations were deduplicated, and subsequently imported into the online tool Rayyan^2^ for screening. Search blocks and filters for PubMed, Embase, and iSearch are detailed below.

PubMed

**Date searched:** 7/5/2021

**Number of results:** 2,884

**Date filter:** January 1, 2020 to present

**Other filters applied:** Language = English

Search blocks

1. covid-19[tw] OR COVID19[tw] OR SARS-CoV-2[tw] OR SARS-CoV2[tw] OR severe acute respiratory syndrome coronavirus 2[tw] OR 2019-nCoV[tw] OR 2019nCoV[tw] OR coronavirus[tw] OR coronavirus[mh] OR covid-19[mh] OR covid[tw]
2. “long COVID”[tw] OR “long covid-19”[tw] OR “long-covid”[tw] OR “long-covid-19”[tw] OR “long haul”[tw] OR “long hauler”[tw] OR “long haulers”[tw] OR “long-haul”[tw] OR “long-hauler”[tw] OR “long-haulers”[tw] OR “chronic COVID”[tw] OR “chronic covid-19”[tw] OR “post-acute COVID”[tw] OR “post-acute covid-19”[tw] OR “post acute COVID”[tw] OR “post acute covid-19”[tw] OR “persistent COVID”[tw] OR “persistent covid-19”[tw] OR “post-COVID”[tw] OR “post-covid-19”[tw] OR “post COVID”[tw] OR “post covid-19”[tw] OR “sequela”[tw] OR “sequelae”[tw] OR “long-term”[tw] OR “long term”[tw] OR “covid syndrome”[tw] OR “covid-19 syndrome”[tw] OR “post-acute COVID-19 syndrome” [Supplementary Concept] OR “persistent symptom”[tw] OR “persistent symptoms”[tw] OR “PASC”[tw] OR “PACS”[tw] OR “PPCS”[tw] OR “post-acute”[tw] OR “post acute”[tw]
3. “prevalent”[tw] OR “prevalence”[tw] OR “prevalence”[mh] OR “occurrence”[tw] OR “occurrences”[tw] OR “duration”[tw] OR “durations”[tw] OR “length”[tw] OR “lengths”[tw] OR “risk factor”[tw] OR “risk factors”[tw] OR (“risk”[tw] AND “factor”[tw]) OR (“risk”[tw] AND “factors”[tw])OR “Risk Factors”[mh] OR “predict”[tw] OR “prediction”[tw] OR “predictions”[tw] OR “predicting”[tw] OR “predictive”[tw] OR “predictor”[tw] OR “predictors”[tw] OR “symptom”[tw] OR “symptoms”[tw] OR “define”[tw] OR “defining”[tw] OR “definition”[tw] OR “definitions”[tw] OR “follow up”[tw] OR “follow-up”[tw] OR “followed up”[tw]

1 AND 2 AND 3

Embase

**Date searched:** 7/5/2021

**Number of results:** 1,390

**Date filter:** January 1, 2020 to present

**Other filters applied:** Language = English, Embase ONLY

1. **‘covid 19’**:ti,ab,de,tn OR **covid19**:ti,ab,de,tn OR **‘sars cov 2’**:ti,ab,de,tn OR **‘sars cov2’**:ti,ab,de,tn OR **‘severe acute respiratory syndrome coronavirus 2’**:ti,ab,de,tn OR **‘2019 ncov’**:ti,ab,de,tn OR **2019ncov**:ti,ab,de,tn OR **coronavirus**:ti,ab,de,tn OR **‘coronavirinae’**/exp OR **‘coronavirus disease 2019’**/exp OR **covid**:ti,ab,de,tn
2. **‘long covid-19’**:ti,ab,de,tn OR **‘long covid’**:ti,ab,de,tn OR **‘long covid 19’**:ti,ab,de,tn OR **‘long haul’**:ti,ab,de,tn OR **‘long hauler’**:ti,ab,de,tn OR **‘long haulers’**:ti,ab,de,tn OR **‘chronic covid’**:ti,ab,de,tn OR **‘chronic covid-19’**:ti,ab,de,tn OR **‘post-acute covid’**:ti,ab,de,tn OR **‘post-acute covid-19’**:ti,ab,de,tn OR **‘post acute covid’**:ti,ab,de,tn OR **‘post acute covid- 19’**:ti,ab,de,tn OR **‘persistent covid’**:ti,ab,de,tn OR **‘persistent covid-19’**:ti,ab,de,tn OR **‘post covid 19’**:ti,ab,de,tn OR **‘post covid’**:ti,ab,de,tn OR **‘post covid-19’**:ti,ab,de,tn OR **sequela**:ti,ab,de,tn OR **sequelae**:ti,ab,de,tn OR **‘long term’**:ti,ab,de,tn OR **‘covid syndrome’**:ti,ab,de,tn OR **‘covid-19 syndrome’**:ti,ab,de,tn OR **‘persistent symptom’**:ti,ab,de,tn OR **‘persistent symptoms’**:ti,ab,de,tn OR **pasc**:ti,ab,de,tn OR **pacs**:ti,ab,de,tn OR **ppcs**:ti,ab,de,tn OR **‘post acute’**:ti,ab,de,tn
3. **prevalent**:ti,ab,de,tn OR **prevalence**:ti,ab,de,tn OR **‘prevalence’**/exp OR **occurrence**:ti,ab,de,tn OR **occurrences**:ti,ab,de,tn OR **duration**:ti,ab,de,tn OR **durations**:ti,ab,de,tn OR **length**:ti,ab,de,tn OR **lengths**:ti,ab,de,tn OR **‘risk factor’**:ti,ab,de,tn OR **‘risk factors’**:ti,ab,de,tn OR (**risk**:ti,ab,de,tn AND **factor**:ti,ab,de,tn) OR (**risk**:ti,ab,de,tn AND **factors**:ti,ab,de,tn) OR **‘risk factor’**/exp OR **predict**:ti,ab,de,tn OR **prediction**:ti,ab,de,tn OR **predictions**:ti,ab,de,tn OR **predicting**:ti,ab,de,tn OR **predictive**:ti,ab,de,tn OR **predictor**:ti,ab,de,tn OR **predictors**:ti,ab,de,tn OR **symptom**:ti,ab,de,tn OR **symptoms**:ti,ab,de,tn OR **define**:ti,ab,de,tn OR **defining**:ti,ab,de,tn OR **definition**:ti,ab,de,tn OR **definitions**:ti,ab,de,tn OR **‘follow up’**:ti,ab,de,tn OR **‘followed up’**:ti,ab,de,tn

1 AND 2 AND 3

iSearch

**Date searched:** 7/5/2021

**Number of results:** 851

**Date filter:** January 1, 2020 to present

**Other filters applied:** Search only title, abstract, preprints only

1. (“long COVID” OR “long covid-19” OR “long-covid” OR “long-covid-19” OR “long haul” OR “long hauler” OR “long haulers” OR “long-haul” OR “long-hauler” OR “long-haulers” OR “chronic COVID” OR “chronic covid-19” OR “post-acute COVID” OR “post-acute covid-19” OR “post acute COVID” OR “post acute covid-19” OR “persistent COVID” OR “persistent covid-19” OR “post-COVID” OR “post-covid-19” OR “post COVID” OR “post covid-19” OR “sequela” OR “sequelae” OR “long-term” OR “long term” OR “covid syndrome” OR “covid-19 syndrome” OR “persistent symptom” OR “persistent symptoms” OR “PASC” OR “PACS” OR “PPCS” OR “post-acute” OR “post acute”)
2. (“prevalent” OR “prevalence” OR “occurrence” OR “occurrences” OR “duration” OR “durations” OR “length” OR “lengths” OR “risk factor” OR “risk factors” OR (“risk” AND “factor”) OR (“risk” AND “factors”) OR “predict” OR “prediction” OR “predictions” OR “predicting” OR “predictive” OR “predictor” OR “predictors” OR “symptom” OR “symptoms” OR “define” OR “defining” OR “definition” OR “definitions” OR “follow up” OR “follow-up” OR “followed up”)

1 AND 2

**eMethods 2.** Sample size calculation

The inclusion/exclusion of sample size in the main text screening was based on a power calculation for Binomial proportions with the formula below:

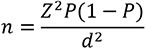

where

n: sample size

Z statistic for 95% CI: z = 1.96, which is the 97.5 percentile point of the standard normal distribution

Expected proportion: P = 0.3

Margin of error: d = 0.05.

**eMethods 3.** Meta-analysis framework

We used random effects model with logit transformation and the DerSimonian-Laird (DL) estimator for *τ*^2^. Thus, the pooled estimated prevalence of PASC 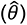 is calculated as:

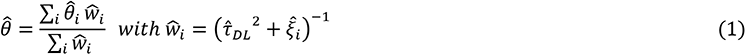

where *i* represents the *i*-th study, 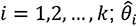 is the logit transformed prevalence *p_i_* in study i so that 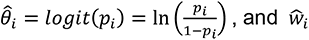 is the estimated weight for study i using the inverse-variance method.

The total variance of study *i* is the sum of within-study variability denoted by 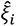, and between-study variability calculated by the DL estimator 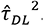. Therefore, 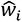 equals to the inverse of the variance, i.e.,

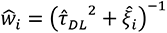

The DL estimator is calculated, as is given below:

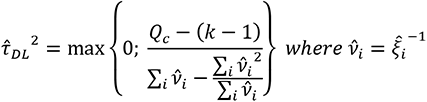

**eMethods 4.** Risk of bias assessment across included articles

Using the Joanna Briggs Institute (JBI) tool^3^, studies were evaluated across 9 common sources of bias in observational studies and given a score out of 9 to reflect how well each study handled these biases. The vast majority (31/40) included studies scored 6/9 or 7/9. Only one study received a perfect score, and our lowest score was 4/9. Supplementary file *Supplementary_RiskofBias.xlsx* contains full results from the risk of bias assessment across the included studies.

Next, we discuss the types of bias that can be introduced in cross-sectional and cohort studies, which make up the majority of the study designs in this review. First, several studies gave COVID-19 cases optional access to a post-COVID clinic or follow-up, which may have resulted in self-selection of sicker individuals. Reporting of symptoms was at times documented through self-report, which has been evidenced to differ from doctor-diagnosed symptoms.^2^ Among studies whose target population was a mixture of hospitalized and non-hospitalized individuals, proportions of hospitalized to non-hospitalized patients varied, possibly biasing the results for this group. Misclassification bias may also be of concern. Patients admitted to the ICU are known to sometimes experience so-called Post-Intensive Care Syndrome (PICS), whose symptomatology is somewhat similar to that of PASC.^3^ Further, some studies suggest that as many as 85% of PASC patients experience symptom resolution, only relapse at a later date, which could obscure the true proportion of individuals experiencing chronic PASC.^4^

---

**Table S1.**
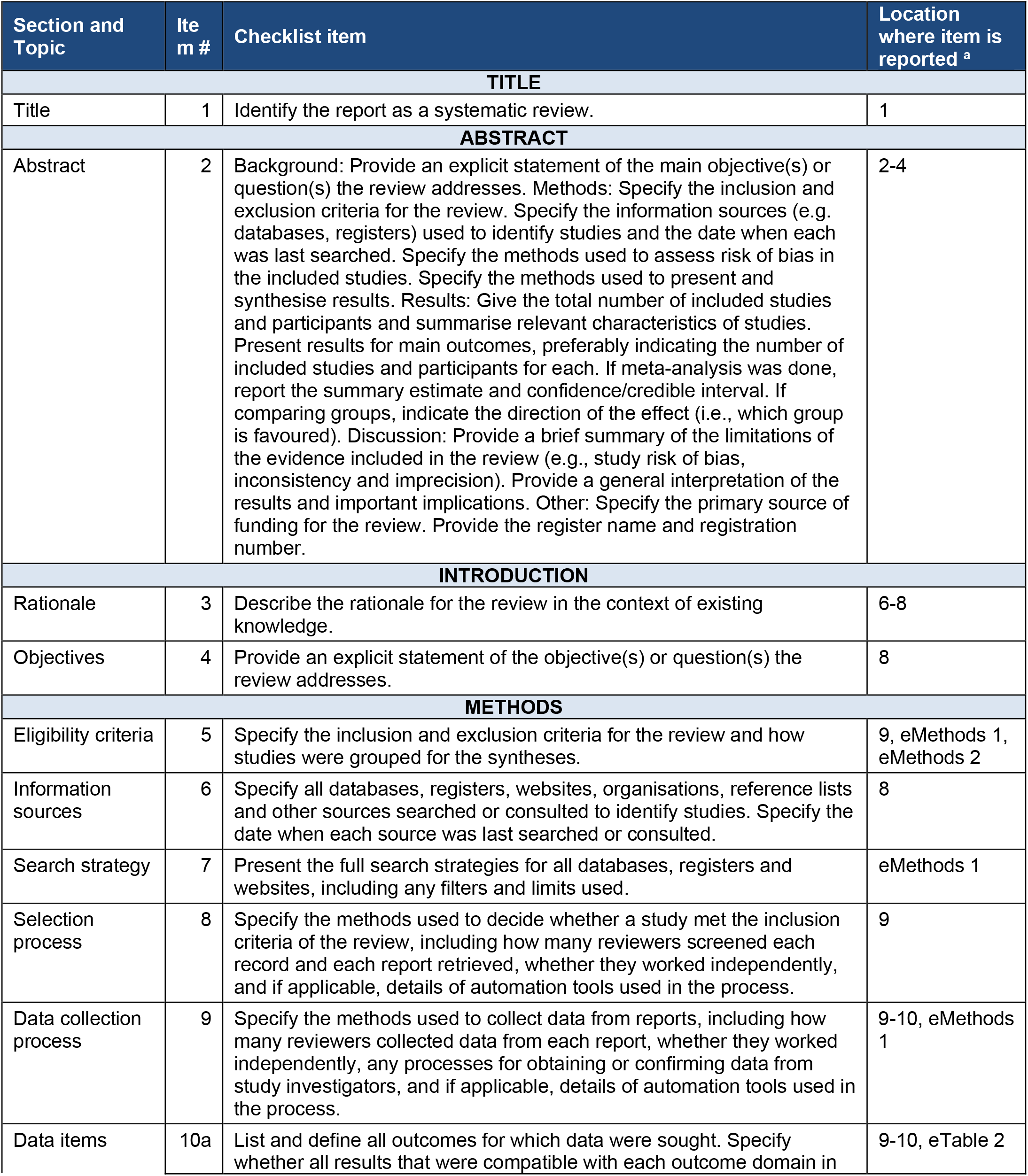
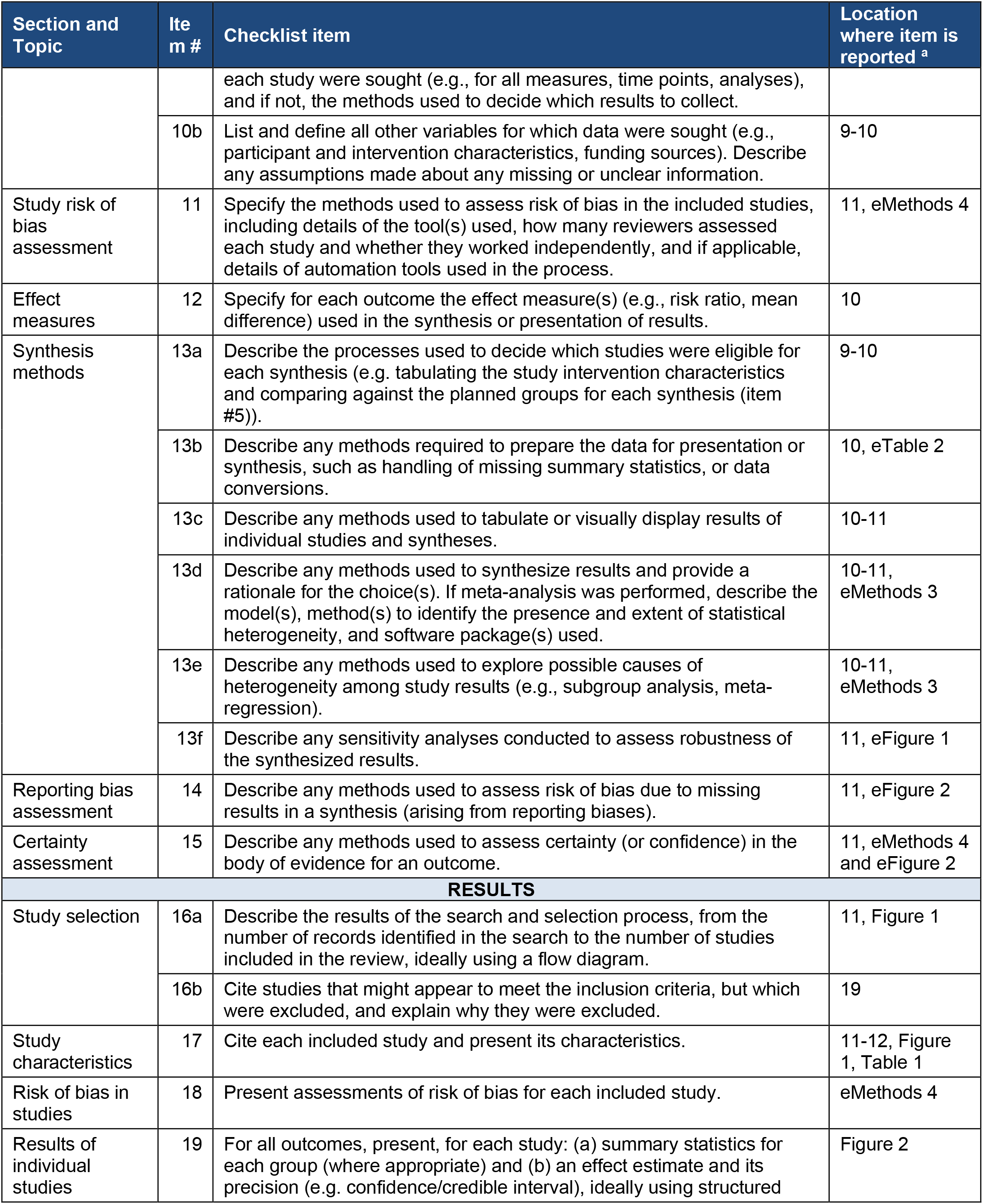
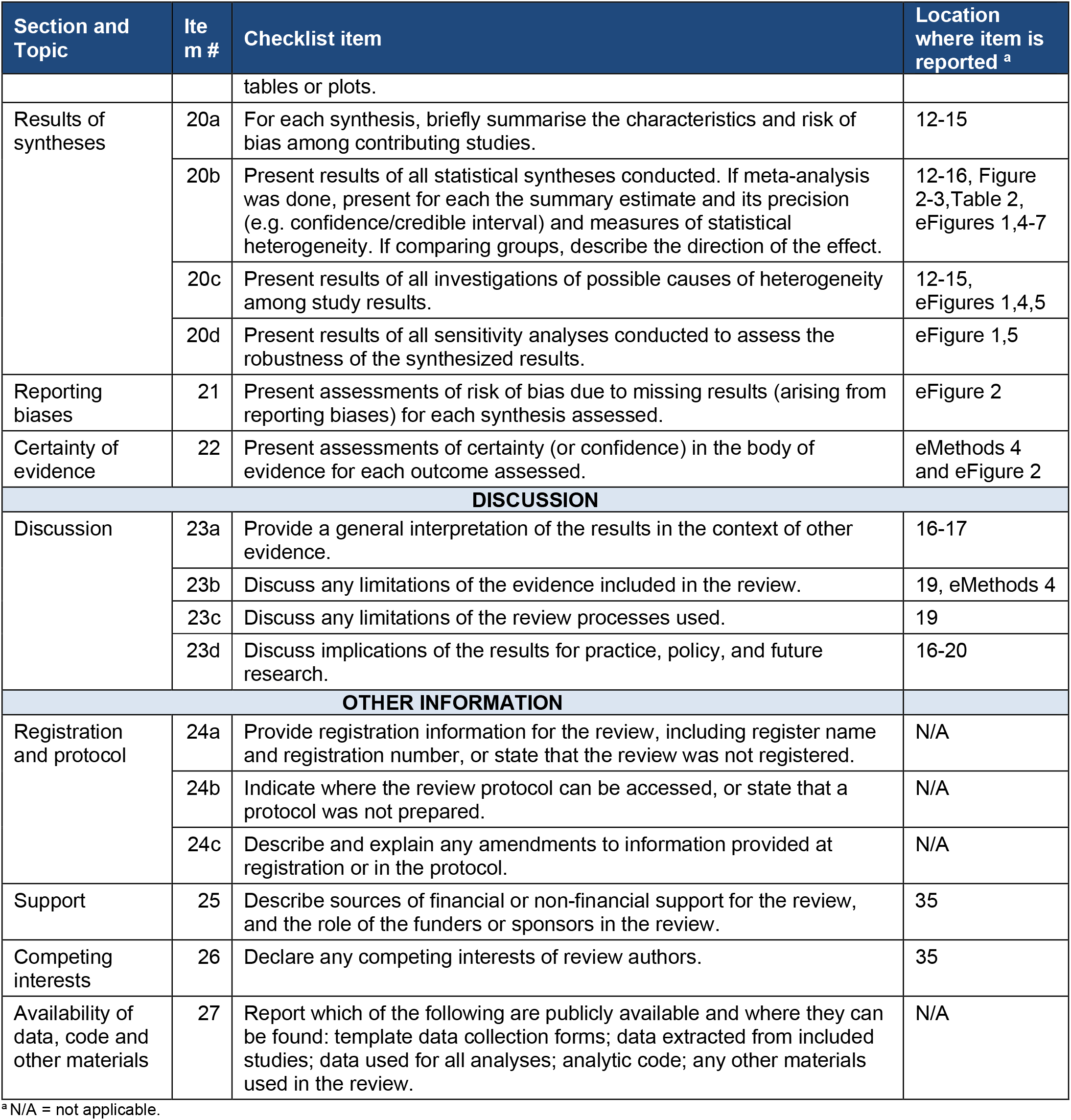
PRISMA checklist

**eTable 1. PRISMA checklist**
| Section and Topic | Item # | Checklist item | Location where item is reported <sup>a</sup> |
| --- | --- | --- | --- |
| **TITLE** |  |  |  |
| Title | 1 | Identify the report as a systematic review. | 1 |
| **ABSTRACT** |  |  |  |
| Abstract | 2 | Background: Provide an explicit statement of the main objective(s) or question(s) the review addresses. Methods: Specify the inclusion and exclusion criteria for the review. Specify the information sources (e.g. databases, registers) used to identify studies and the date when each was last searched. Specify the methods used to assess risk of bias in the included studies. Specify the methods used to present and synthesise results. Results: Give the total number of included studies and participants and summarise relevant characteristics of studies. Present results for main outcomes, preferably indicating the number of included studies and participants for each. If meta-analysis was done, report the summary estimate and confidence/credible interval. If comparing groups, indicate the direction of the effect (i.e., which group is favoured). Discussion: Provide a brief summary of the limitations of the evidence included in the review (e.g., study risk of bias, inconsistency and imprecision). Provide a general interpretation of the results and important implications. Other: Specify the primary source of funding for the review. Provide the register name and registration number. | 2-4 |
| **INTRODUCTION** |  |  |  |
| Rationale | 3 | Describe the rationale for the review in the context of existing knowledge. | 6-8 |
| Objectives | 4 | Provide an explicit statement of the objective(s) or question(s) the review addresses. | 8 |
| **METHODS** |  |  |  |
| Eligibility criteria | 5 | Specify the inclusion and exclusion criteria for the review and how studies were grouped for the syntheses. | 9, eMethods 1, eMethods 2 |
| Information sources | 6 | Specify all databases, registers, websites, organisations, reference lists and other sources searched or consulted to identify studies. Specify the date when each source was last searched or consulted. | 8 |
| Search strategy | 7 | Present the full search strategies for all databases, registers and websites, including any filters and limits used. | eMethods 1 |
| Selection process | 8 | Specify the methods used to decide whether a study met the inclusion criteria of the review, including how many reviewers screened each record and each report retrieved, whether they worked independently, and if applicable, details of automation tools used in the process. | 9 |
| Data collection process | 9 | Specify the methods used to collect data from reports, including how many reviewers collected data from each report, whether they worked independently, any processes for obtaining or confirming data from study investigators, and if applicable, details of automation tools used in the process. | 9-10, eMethods 1 |
| Data items | 10a | List and define all outcomes for which data were sought. Specify whether all results that were compatible with each outcome domain in each study were sought (e.g., for all measures, time points, analyses), and if not, the methods used to decide which results to collect. | 9-10, eTable 2 |
|  | 10b | List and define all other variables for which data were sought (e.g., participant and intervention characteristics, funding sources). Describe any assumptions made about any missing or unclear information. | 9-10 |
| Study risk of bias assessment | 11 | Specify the methods used to assess risk of bias in the included studies, including details of the tool(s) used, how many reviewers assessed each study and whether they worked independently, and if applicable, details of automation tools used in the process. | 11, eMethods 4 |
| Effect measures | 12 | Specify for each outcome the effect measure(s) (e.g., risk ratio, mean difference) used in the synthesis or presentation of results. | 10 |
| Synthesis methods | 13a | Describe the processes used to decide which studies were eligible for each synthesis (e.g. tabulating the study intervention characteristics and comparing against the planned groups for each synthesis (item #5)). | 9-10 |
|  | 13b | Describe any methods required to prepare the data for presentation or synthesis, such as handling of missing summary statistics, or data conversions. | 10, eTable 2 |
|  | 13c | Describe any methods used to tabulate or visually display results of individual studies and syntheses. | 10-11 |
|  | 13d | Describe any methods used to synthesize results and provide a rationale for the choice(s). If meta-analysis was performed, describe the model(s), method(s) to identify the presence and extent of statistical heterogeneity, and software package(s) used. | 10-11, eMethods 3 |
|  | 13e | Describe any methods used to explore possible causes of heterogeneity among study results (e.g., subgroup analysis, meta-regression). | 10-11, eMethods 3 |
|  | 13f | Describe any sensitivity analyses conducted to assess robustness of the synthesized results. | 11, eFigure 1 |
| Reporting bias assessment | 14 | Describe any methods used to assess risk of bias due to missing results in a synthesis (arising from reporting biases). | 11, eFigure 2 |
| Certainty assessment | 15 | Describe any methods used to assess certainty (or confidence) in the body of evidence for an outcome. | 11, eMethods 4 and eFigure 2 |
| **RESULTS** |  |  |  |
| Study selection | 16a | Describe the results of the search and selection process, from the number of records identified in the search to the number of studies included in the review, ideally using a flow diagram. | 11, Figure 1 |
|  | 16b | Cite studies that might appear to meet the inclusion criteria, but which were excluded, and explain why they were excluded. | 19 |
| Study characteristics | 17 | Cite each included study and present its characteristics. | 11-12, Figure 1, Table 1 |
| Risk of bias in studies | 18 | Present assessments of risk of bias for each included study. | eMethods 4 |
| Results of individual studies | 19 | For all outcomes, present, for each study: (a) summary statistics for each group (where appropriate) and (b) an effect estimate and its precision (e.g. confidence/credible interval), ideally using structured tables or plots. | Figure 2 |
| Results of syntheses | 20a | For each synthesis, briefly summarise the characteristics and risk of bias among contributing studies. | 12-15 |
|  | 20b | Present results of all statistical syntheses conducted. If meta-analysis was done, present for each the summary estimate and its precision (e.g. confidence/credible interval) and measures of statistical heterogeneity. If comparing groups, describe the direction of the effect. | 12-16, Figure 2-3, Table 2, eFigures 1,4-7 |
|  | 20c | Present results of all investigations of possible causes of heterogeneity among study results. | 12-15, eFigures 1,4,5 |
|  | 20d | Present results of all sensitivity analyses conducted to assess the robustness of the synthesized results. | eFigure 1,5 |
| Reporting biases | 21 | Present assessments of risk of bias due to missing results (arising from reporting biases) for each synthesis assessed. | eFigure 2 |
| Certainty of evidence | 22 | Present assessments of certainty (or confidence) in the body of evidence for each outcome assessed. | eMethods 4 and eFigure 2 |
| **DISCUSSION** |  |  |  |
| Discussion | 23a | Provide a general interpretation of the results in the context of other evidence. | 16-17 |
|  | 23b | Discuss any limitations of the evidence included in the review. | 19, eMethods 4 |
|  | 23c | Discuss any limitations of the review processes used. | 19 |
|  | 23d | Discuss implications of the results for practice, policy, and future research. | 16-20 |
| **OTHER INFORMATION** |  |  |  |
| Registration and protocol | 24a | Provide registration information for the review, including register name and registration number, or state that the review was not registered. | N/A |
|  | 24b | Indicate where the review protocol can be accessed, or state that a protocol was not prepared. | N/A |
|  | 24c | Describe and explain any amendments to information provided at registration or in the protocol. | N/A |
| Support | 25 | Describe sources of financial or non-financial support for the review, and the role of the funders or sponsors in the review. | 35 |
| Competing interests | 26 | Declare any competing interests of review authors. | 35 |
| Availability of data, code and other materials | 27 | Report which of the following are publicly available and where they can be found: template data collection forms; data extracted from included studies; data used for all analyses; analytic code; any other materials used in the review. | N/A |

<sup>a</sup> N/A = not applicable.

**Table S2.**
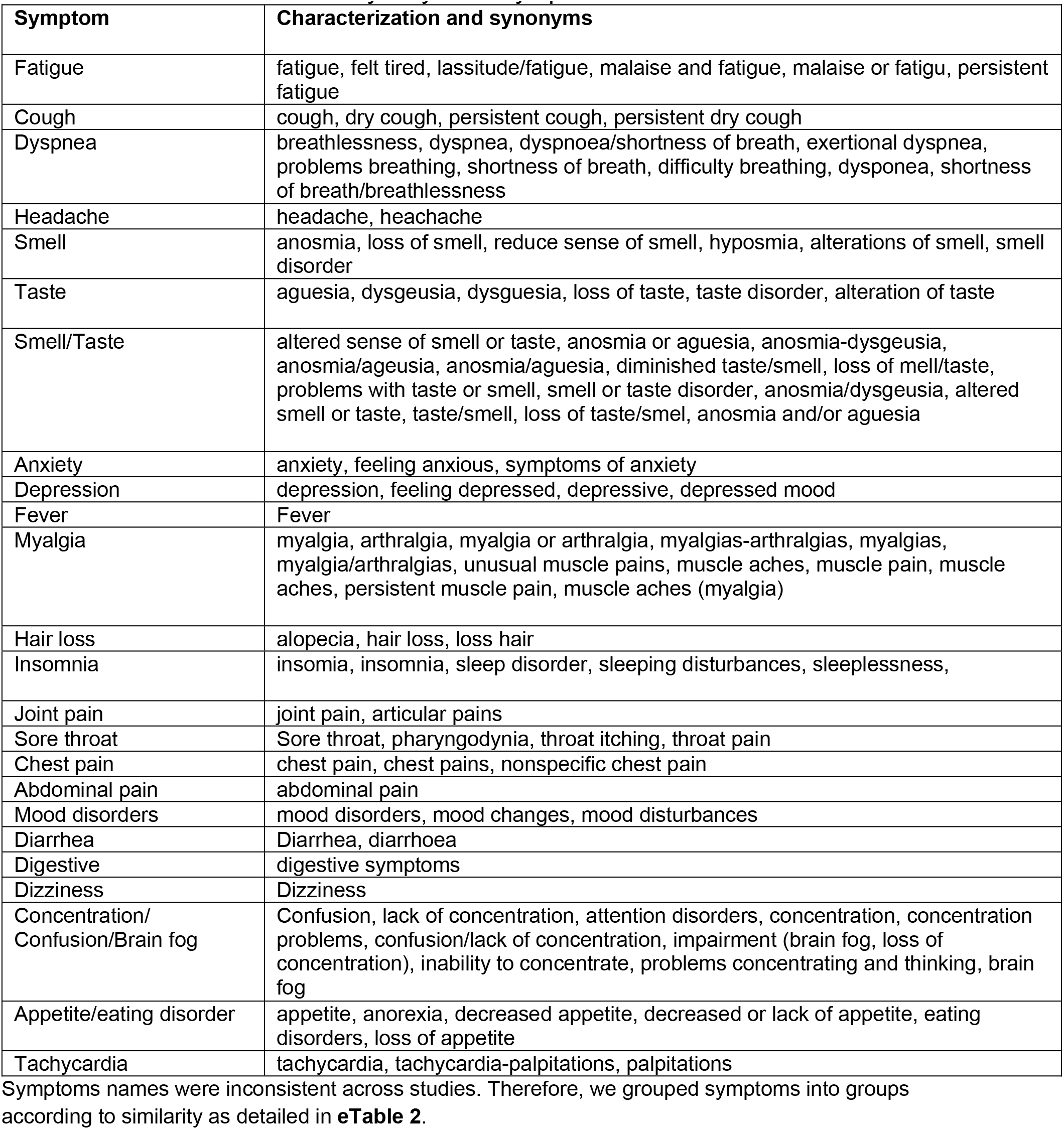
Characterization and synonyms of symptoms

**Table S3.** Summary of excluded articles by region and follow-up time ^a, b^

| **Exclusion Reason** | **Africa** | **Asia** | **Australia** | **Europe** | **North America** | **South America** | **Mix** | **Total** |
| --- | --- | --- | --- | --- | --- | --- | --- | --- |
| Abstract only |  |  | 1 | 8 | 8 |  |  | 17 |
| Duplicate cohort |  |  |  | 1 |  |  |  | 1 |
| Duplicated |  |  |  |  |  |  |  | 0 |
| Wrong Follow-up Time | 4 | 6 |  | 9 | 2 |  | 1 | 22 |
| Wrong Index Date | 1 |  |  | 1 |  |  |  | 2 |
| Sample Size | 2 | 7 | 1 | 38 | 11 | 1 | 1 | 61 |
| Wrong Outcome |  | 8 |  | 28 | 16 |  | 4 | 56 |
| Wrong Population |  | 1 |  | 14 | 6 | 1 | 4 | 26 |
| Wrong Publication Type |  |  |  | 4 | 1 |  |  | 5 |
| Total | 7 | 22 | 2 | 103 | 44 | 2 | 10 | 190 |

<sup>a</sup> Studies excluded during the full-text screening are represented in this table.<sup>b</sup> Studies where region is not applicable or unknown are not represented in this table

**Table S4.** Summary of significant non-meta-analyzed risk factors for PASC

**eTable 4.** Summary of significant non-meta-analyzed risk factors for PASC
| **Studies** | **Predictor** | **Odds Ratio (OR)** | **95% lower confidence bound** | **95% upper confidence bound** |
| --- | --- | --- | --- | --- |
| Augustin et al.<sup>5</sup> | Number of symptoms in acute COVID-19 | 1.29 | 1.08 | 1.55 |
| Naik et al.<sup>6</sup> | Hypothyroidism | 4.13 | 2.20 | 7.60 |
|  | COVID-19 severity: moderate vs mild | 1.70 | 1.10 | 2.40 |
| Menges et al.<sup>7</sup> | Severe to very severe initial symptoms (mild to moderate reference group) | 2.05 | 1.27 | 3.34 |
|  | Comorbidities | 2.08 | 1.24 | 3.50 |
| Perlis et al.<sup>8</sup> | Covid severity: somewhat vs not at all | 2.59 | 1.75 | 3.94 |
|  | Covid severity: very vs not at all | 3.78 | 2.54 | 5.79 |
|  | Muscle pain | 1.31 | 1.03 | 1.67 |
|  | Headache | 1.44 | 1.11 | 1.86 |
|  | Shaking | 1.3 | 1.01 | 1.66 |
|  | Age (decade) | 1.1 | 1.02 | 1.2 |
| Yomogida et al.<sup>10</sup> | Pre-existing condition | 2.14 | 1.33 | 3.43 |
|  | Asthma | 4.50 | 2.12 | 9.57 |
|  | Obesity | 7.73 | 2.51 | 23.83 |
|  | Hypertension | 1.86 | 1.09 | 3.18 |
|  | Severe pre-existing conditions (stroke, cancer, immunocompromising conditions, liver, lung, and kidney disorders) | 2.46 | 1.14 | 5.30 |
|  | Severe Covid at diagnosis (asymptomatic reference group) | 13.33 | 2.43 | 73.02 |
|  | Moderate Covid at diagnosis (asymptomatic reference group) | 12.75 | 2.86 | 56.88 |

| Studies | Predictor | Odds Ratio (OR) | 95% lower confidence bound | 95% upper confidence bound |
| --- | --- | --- | --- | --- |
|  | Mild Covid at diagnosis (asymptomatic reference group) | 5.00 | 1.51 | 21.77 |
|  | Age 40-54 (25-39 reference group) | 1.81 | 1.05 | 3.12 |
| Sudre et al.<sup>11</sup> | Fatigue in first week | 2.83 | 2.09 | 3.83 |
|  | Headache in first week | 2.62 | 2.04 | 3.37 |
|  | Dyspnea in first week | 2.36 | 1.91 | 2.91 |
|  | Hoarse voice in first week | 2.33 | 1.88 | 2.90 |
|  | Myalgia in first week | 2.22 | 1.80 | 2.73 |
| Desgranges<sup>12</sup> | Female | 1.67 | 1.09 | 2.56 |
|  | Overweight/obese | 1.67 | 1.10 | 2.56 |

**eTable 4** presents a list of non-meta-analyzed risk factors and their associated odds ratios and confidence intervals. Only factors with a significant positive association with PASC were included (i.e., OR > 1 and CI does not cross 1). If both odds ratios and adjusted odds ratios were provided for a risk factor, we used the adjusted odds ratio.

**Table S5.** Summary of duration of PASC and symptoms

| Outcome | Median duration of PASC or symptoms by studies [IQR] |  |  |
| --- | --- | --- | --- |
|  | *Sudre et al.*<sup>11</sup> |  | *Naik et al.*<sup>6</sup> |
|  | *28+ days* | *56+ days* | *28 days+* |
| PASC | 41 |  |  |
| Abdominal pain | 7 | 13 |  |
| Chest pain | 13 | 46 | 60 [41-112] |
| Cough | 20 | 34 | 60 [45-118] |
| Delirium | 8 | 14 |  |
| Dyspnea | 24 | 59 | 90 [45-124] |
| Fatigue | 33 | 73 | 60 [45-135] |
| Headache | 22 | 56 |  |
| Myalgia | 7 | 30 | 60 [45-150] |
| Loss of smell | 24 | 53 |  |
| Sore throat | 15 | 33 |  |
| Fever | 6 | 11 |  |
| Disturbed sleep |  |  | 45 [40-70] |

Summary of two studies that evaluated median duration of PASC and specific symptoms. Sudre et al further calculated median duration by different follow-up time, at least 28 days after index date and at least 56 days after the index date.

**Figure S1.**
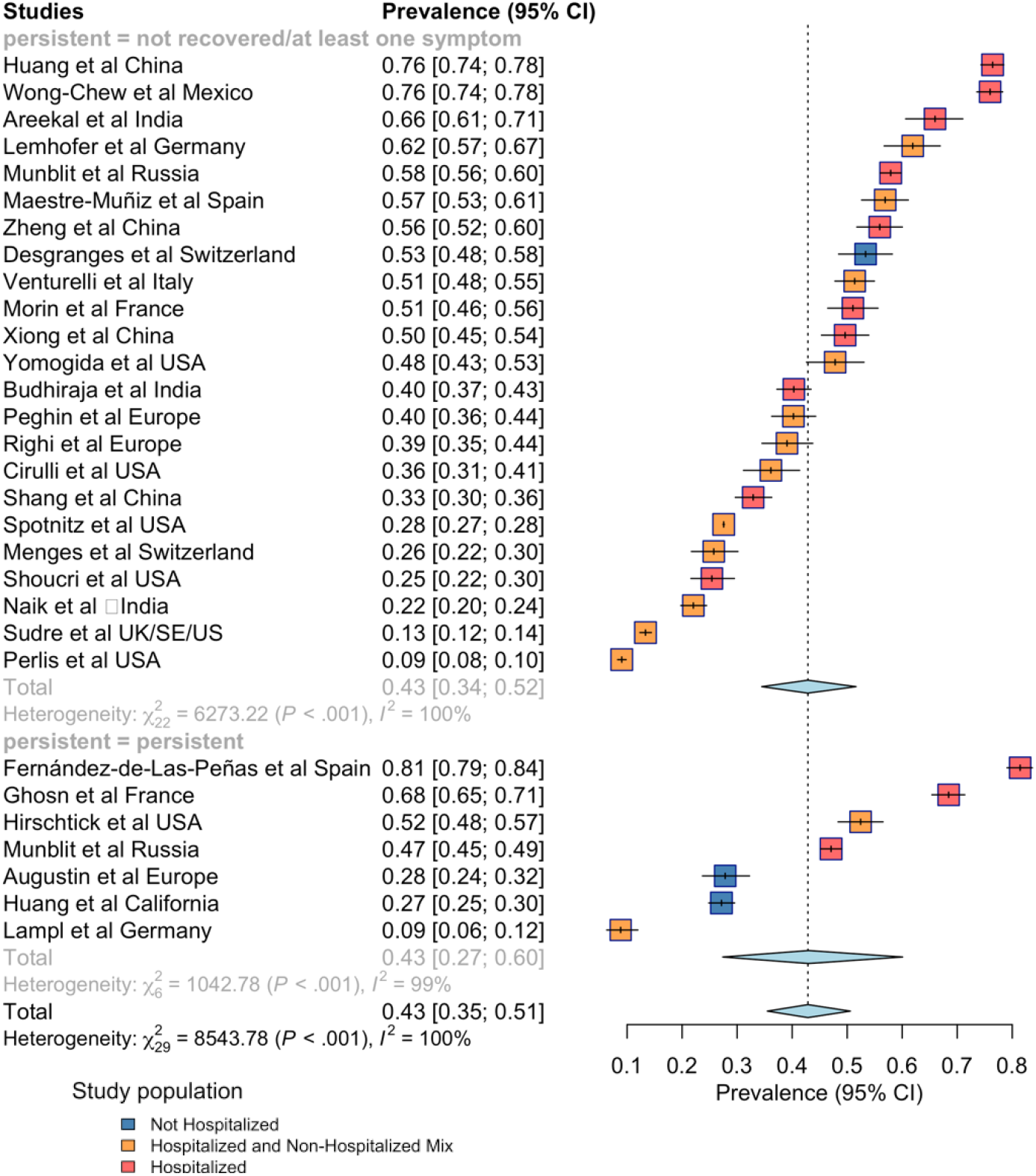
Stratified analysis of PASC by persisted versus not persisted symptoms PASC definitions varied across studies. **eFigure 1** stratified PASC prevalence according to whether a study defined PASC as having at least one symptom or not recovered at follow-up, or having persistent symptoms at follow-up.

**Figure S2.**
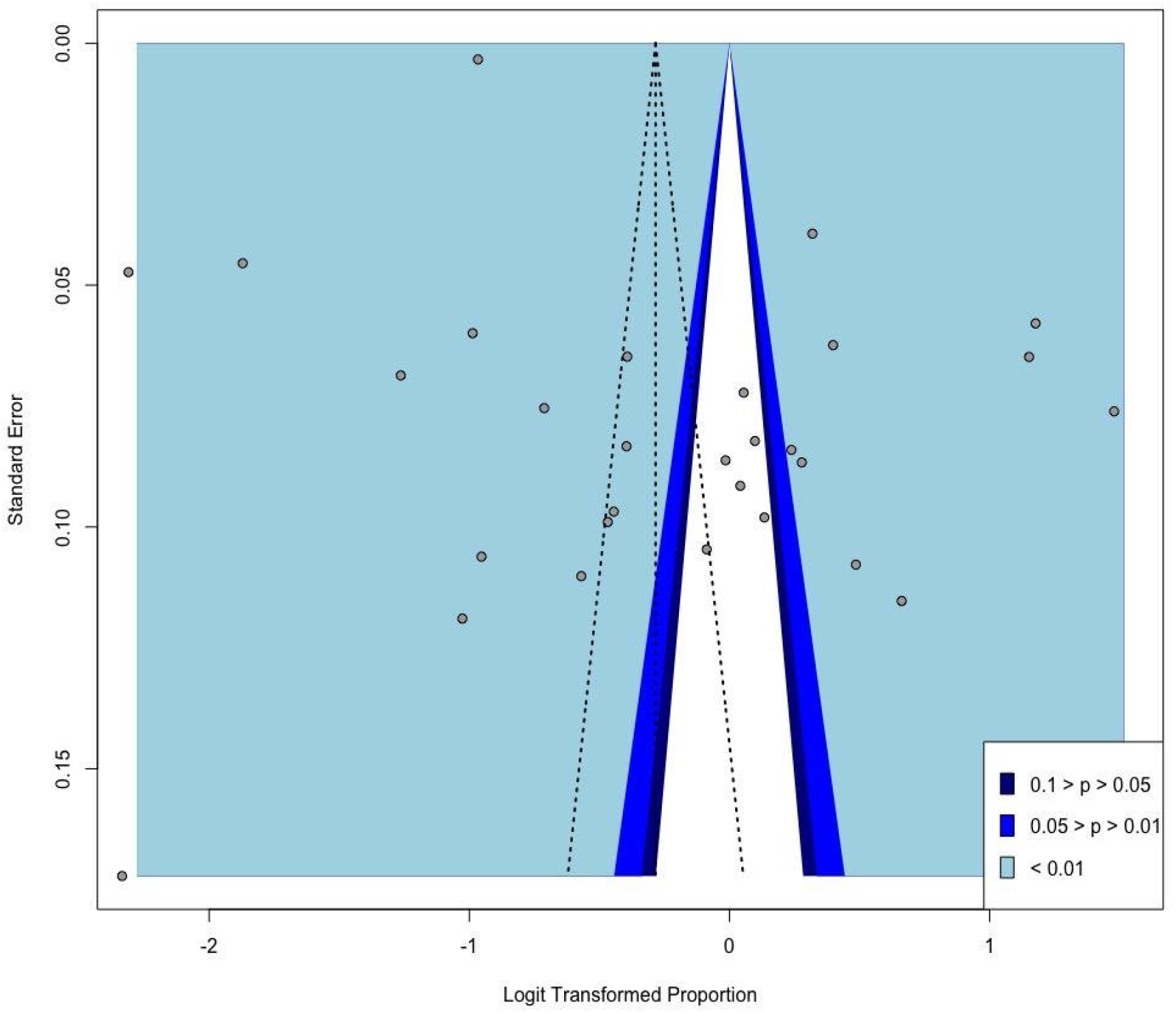
Funnel plot for publication bias assessment among included studies **eFigure 2** presents the funnel plot for examination of publication bias among the included studies in the quantitative synthesis. The standard procedure of an initial visual inspection for publication bias suggests that asymmetry may be present. In further formally testing for presence of asymmetry, the Egger’s test statistic of 3.18 is significant (p-value of 0.003 < 0.05), and as a further check, the Begg rank correlation test is performed with a resulting test statistic that is not significant (p-value of 0.679 > 0.05). The contrasting results is generally not uncommon, as the concordance between these two tests has been found to be moderate.^4^ Moreover, the Begg test has been evidenced to result in larger p-values for meta-analyses without a considerably number of studies of which the synthesis herein may qualify (i.e., number of included studies is 29; threshold for small meta-analyses is 25 studies, whereas roughly 75 studies constitute a large meta-analysis).^13^ In sum, although the Egger’s test indicates a detection of bias from publishing, for meta-analyzing proportion estimates from observational studies, asymmetry in this context does not strictly indicate publication bias as proportions are published without preference over a particular effect size^5^. Additionally, with most included articles being hospital-based or multi-center study designs, the results may be driven by the larger sample sizes. Additionally, we suspect that heterogeneity in the true effect sizes of PASC prevalence, across the geographic entities reflected in the included studies, accounts for some of the horizontal spread in the proportion estimates ^5^.

**Figure S3.**
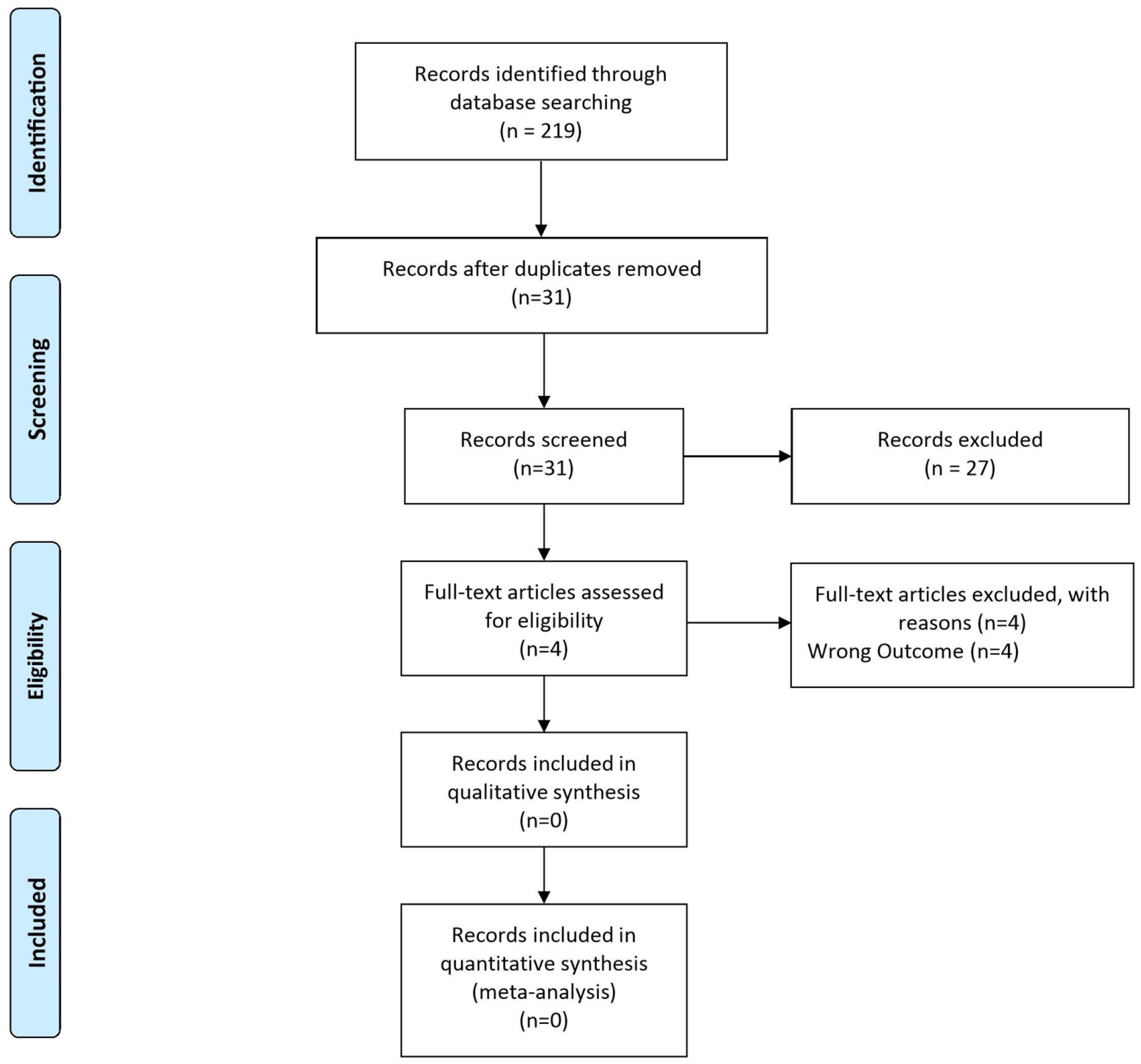
PRISMA flow diagram for extended search **eFigure 3** details the study selection process for the search extension we performed on August 12, 2021 as described in Supplement B. We found 31 new studies (not covered in the original search) from selected journals, which we then screened. None of these studies met our inclusion/exclusion criteria, and thus none were included in the systematic review or meta-analysis.

**Figure S4.**
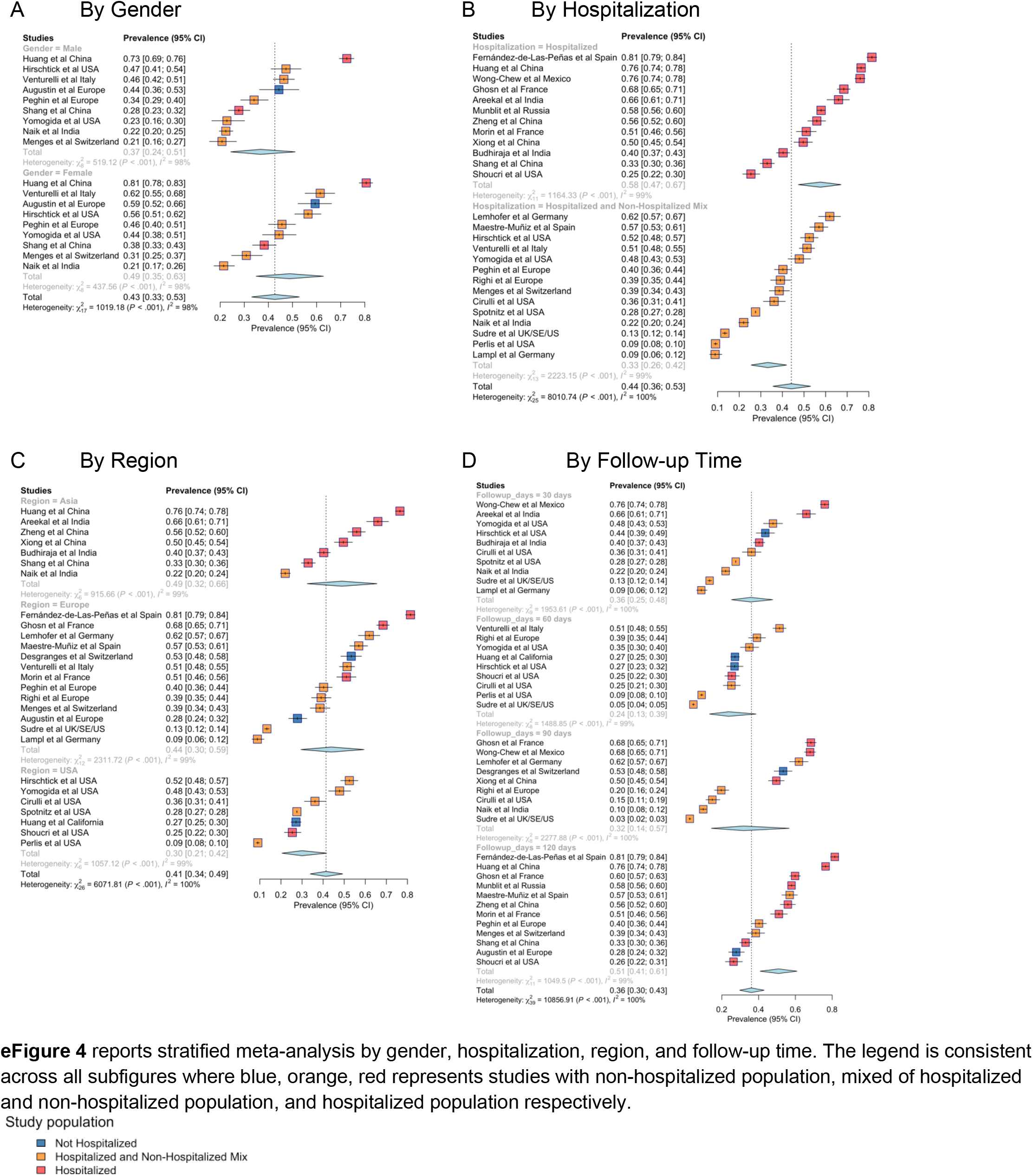
Forest plots with the prevalence estimates of PASC stratified by (A) gender (B) hospitalization, (C) region, and (D) follow-up time, respectively

**Figure S5.**
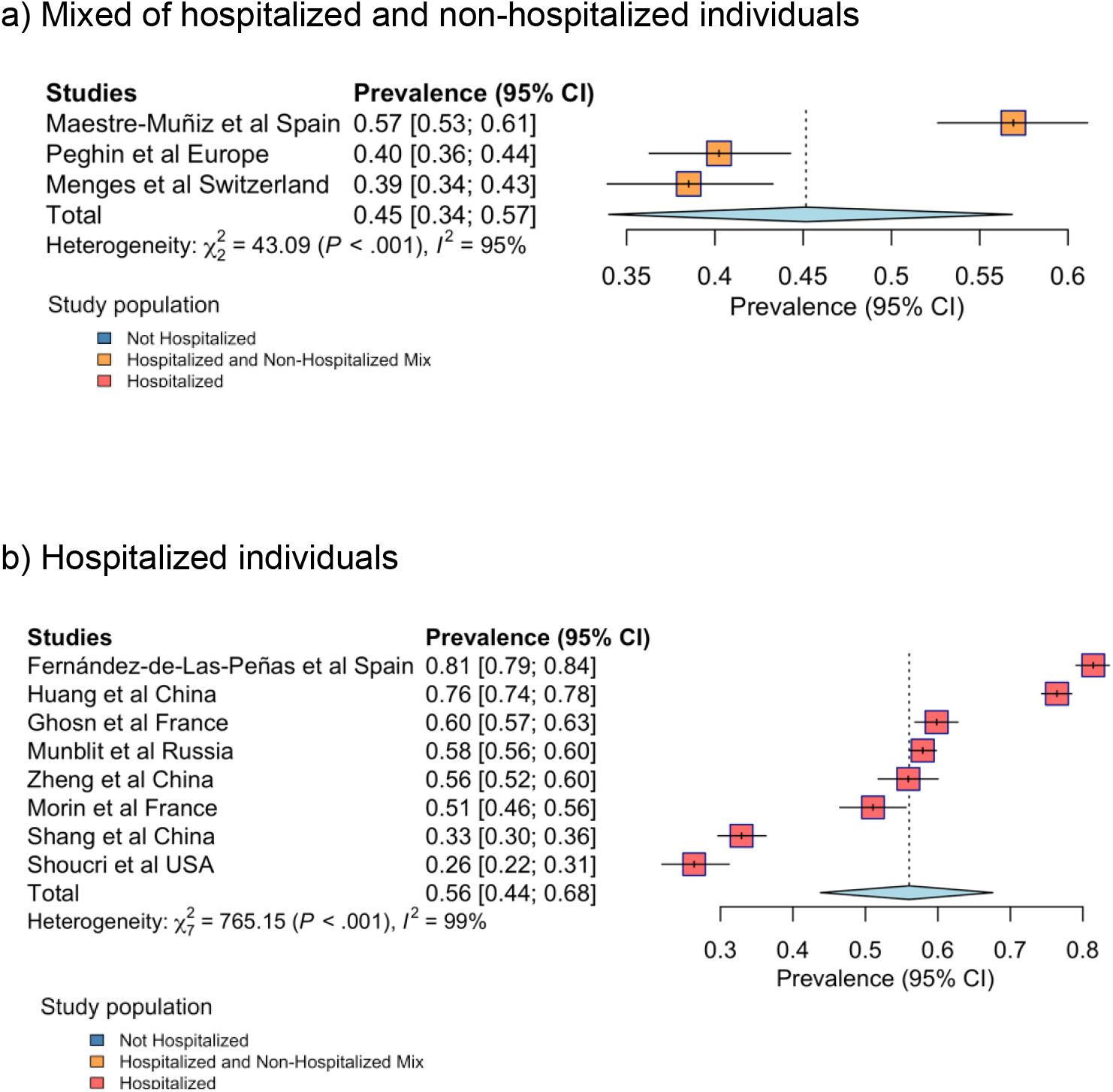
Meta-analysis of studies with 120 follow-up days stratified by acute-phase hospitalization status of study population

**Figure S6.**
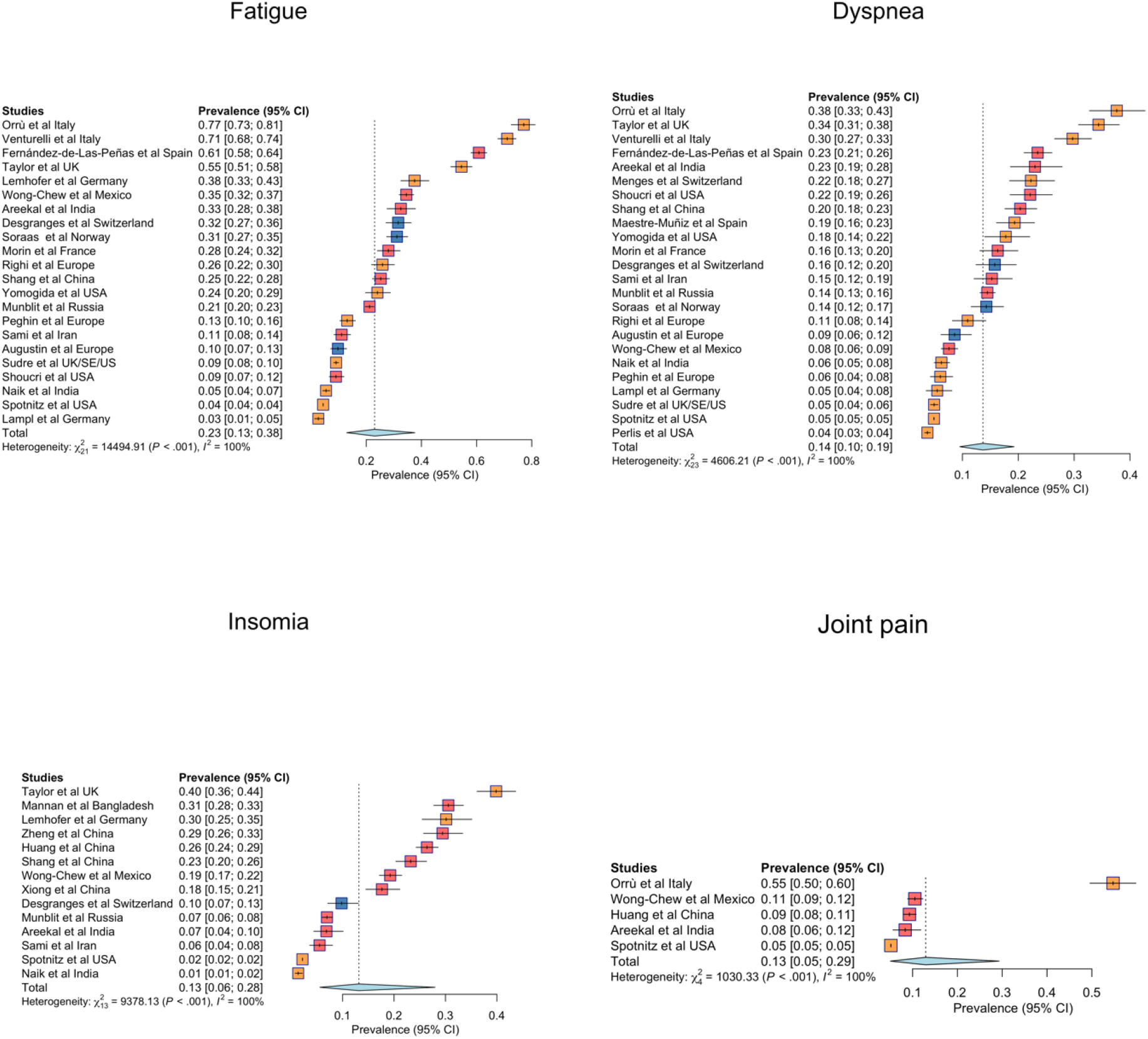
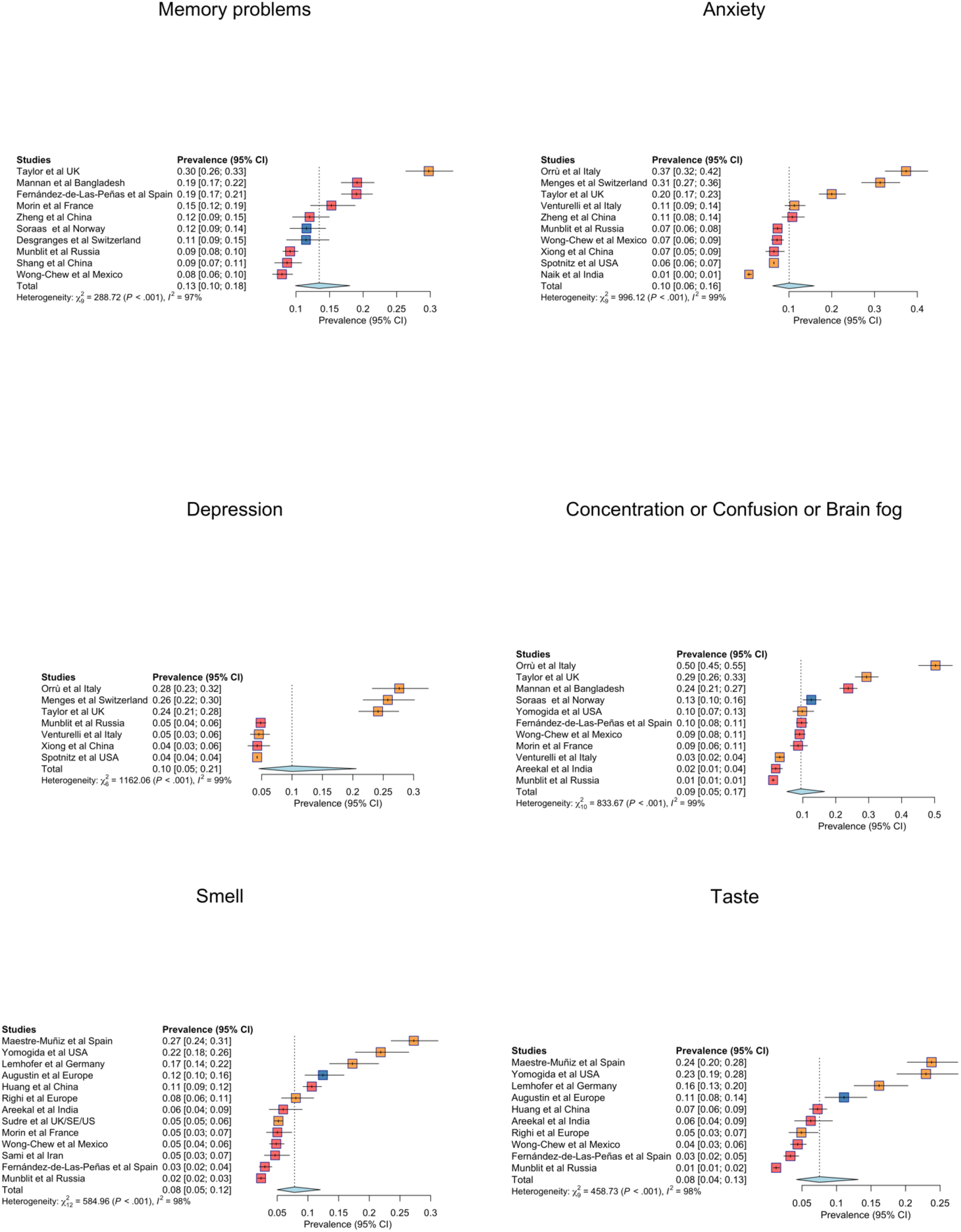
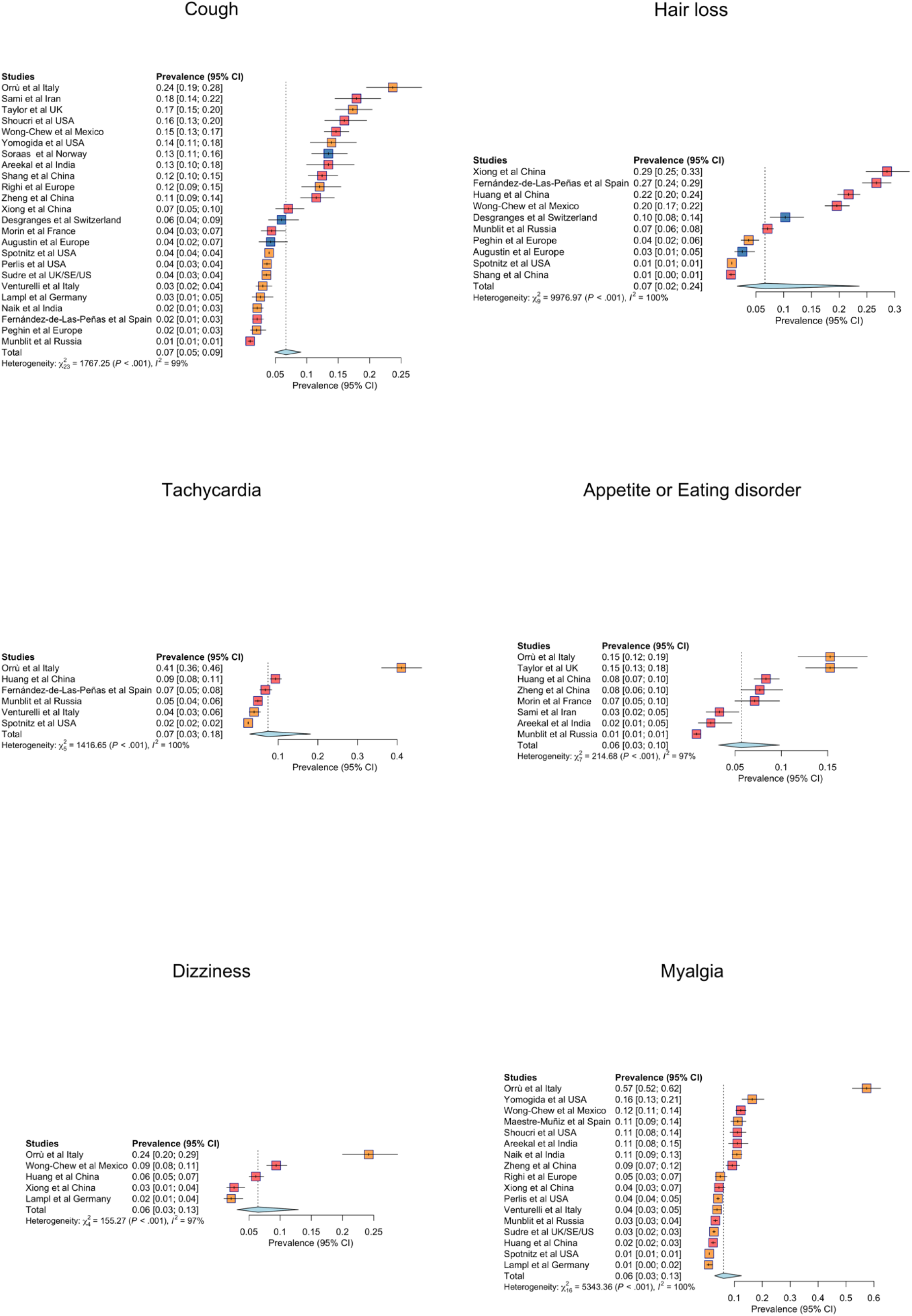
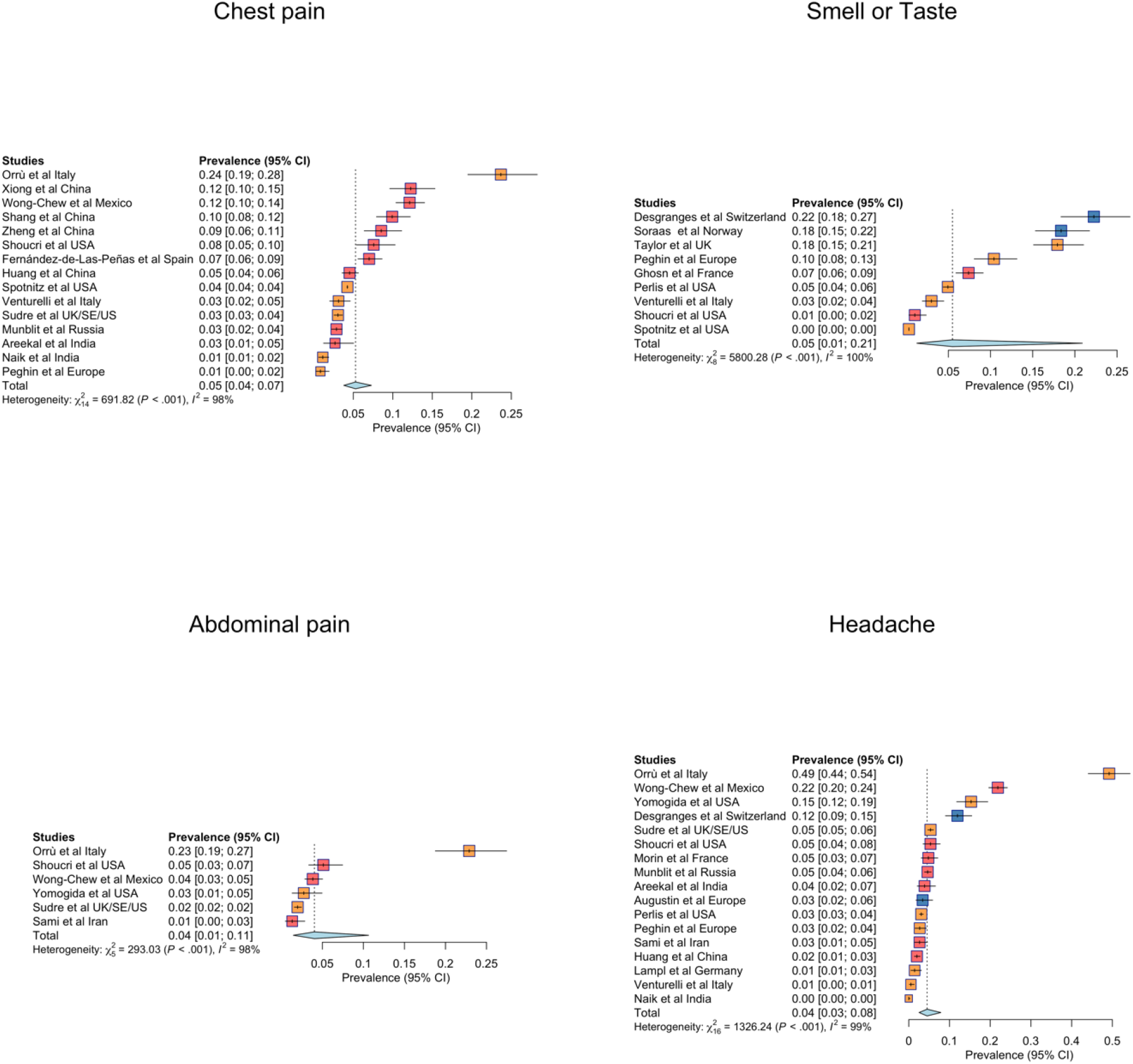
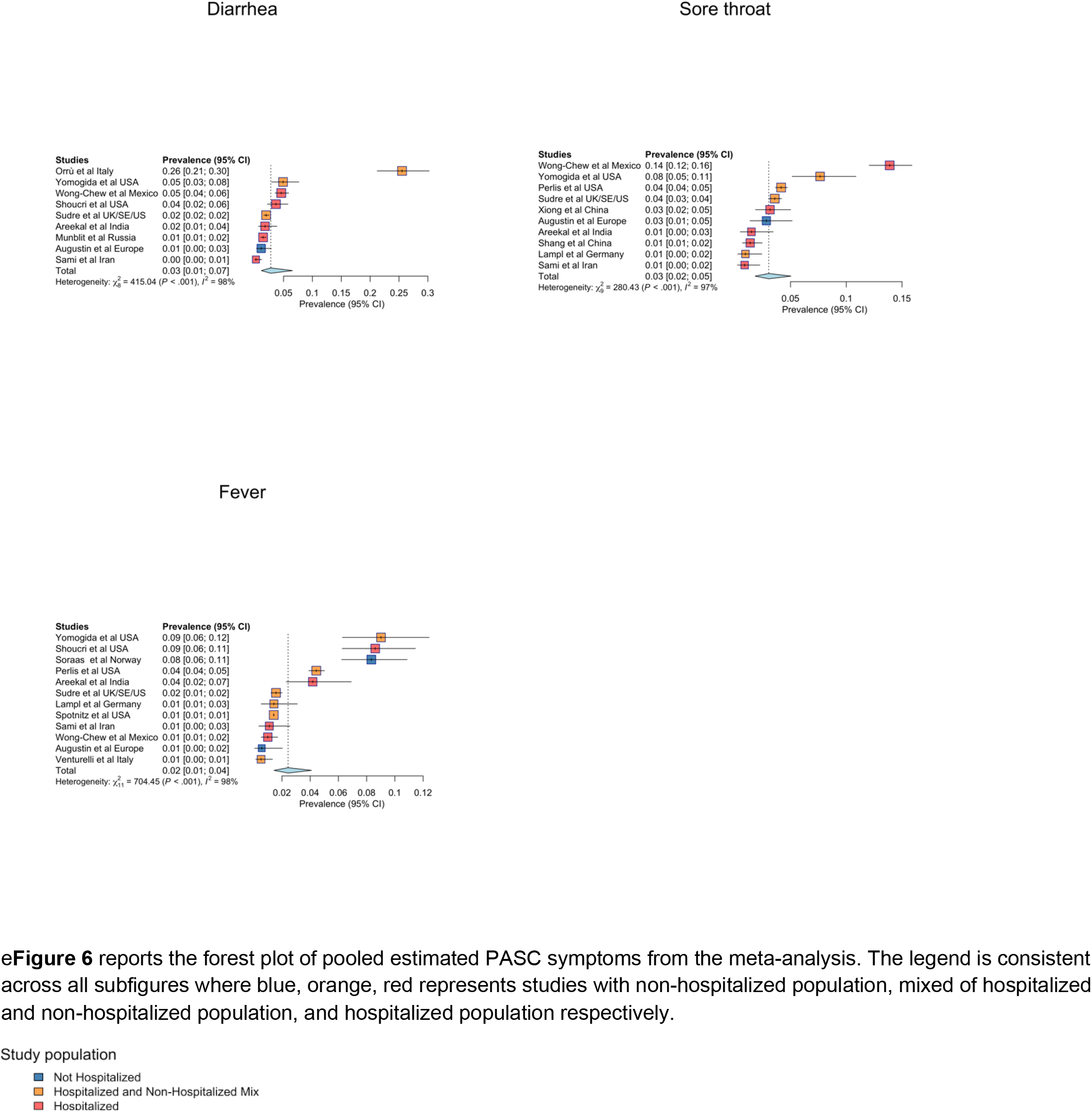
Supplementary meta-analysis of PASC symptom-specific prevalence

**Figure S7.**
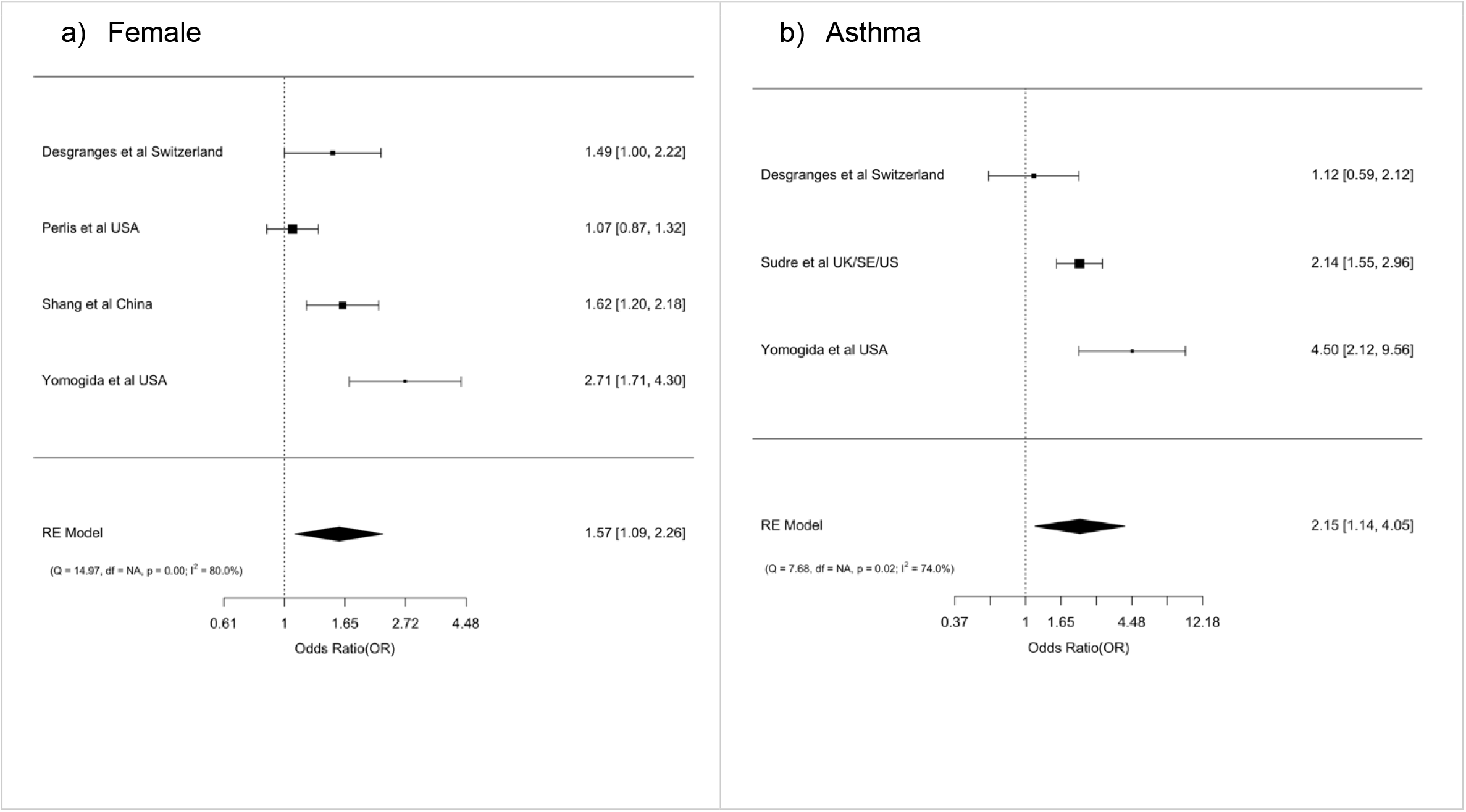
Forest plots of risk factors for PASC by a) female and b) asthma **Figure S7** reports estimated Odds Ratios for female sex and pre-existing asthma as a predictor of PASC.

